# Mendelian randomization of dyslipidemia on cognitive impairment among older Americans

**DOI:** 10.1101/2020.10.20.20216036

**Authors:** Mingzhou Fu, Kelly M. Bakulski, Cesar Higgins, Erin B. Ware

## Abstract

**Background:** Altered lipid metabolism may be a risk factor for dementia, and blood cholesterol level has a strong genetic component. We tested the hypothesis that dyslipidemia (either low levels of high-density lipoprotein cholesterol (HDL-C) or high total cholesterol) is associated with cognitive status and domains, and assessed inferred causality using genetic predisposition to dyslipidemia as an instrumental variable.

**Methods:** Using data from European and African genetic ancestry participants in the Health and Retirement Study, we selected observations at the first non-missing biomarker assessment (waves 2006 to 2012). Cognition domains were assessed using episodic memory, mental status, and vocabulary tests. Overall cognitive status was categorized in three levels (normal, cognitive impairment non-dementia, dementia). Based on 2018 clinical guidelines, we compared low HDL-C or high total cholesterol to normal levels. Polygenic scores for dyslipidemia were used as instrumental variables in a Mendelian randomization framework. Multivariable logistic regressions and Wald-type ratio estimators were used to examine associations.

**Results:** Among European ancestry participants (n = 8781), at risk HDL-C levels were associated with higher odds of cognitive impairment (OR = 1.20, 95%CI: 1.03, 1.40) and worse episodic memory, specifically. Using cumulative genetic risk for HDL-C levels as a valid instrumental variable, a significant causal estimate was observed between at risk low HDL-C levels and higher odds of dementia (OR = 2.15, 95%CI: 1.16, 3.99). No significant associations were observed between total cholesterol levels and cognitive status. No significant associations were observed in the African ancestry sample (n = 2101).

**Conclusion:** Our study demonstrates low blood HDL-C is a potential causal risk factor for impaired cognition during aging in non-Hispanic whites of European ancestry. Dyslipidemia can be modified by changing diets, health behaviors, and therapeutic strategies, which can improve cognitive aging. Studies on low density lipoprotein cholesterol, the timing of cholesterol effects on cognition, and larger studies in non-European ancestries are needed.

## Background

Dementia is a chronic and progressive syndrome beyond normal cognitive decline in aging. It affects memory, mental status, and activities of daily living, resulting in disability and dependency. There is no treatment currently available to cure or to alter the progressive course of dementia.^1^ Worldwide, over 50 million people are currently living with dementia, with 10 million new cases every year.^1^ The cost for global dementia treatment is expected to reach two trillion United States dollars by 2030.^2^

Altered lipid metabolism is implicated in dementia pathogenesis. Several epidemiological studies show associations between fundamental differences in cholesterol levels and cognitive decline risks. A meta-analysis reported a relative risk of 2.14 for participants with high total cholesterol (TC) in midlife to develop all-type dementia compared to participants with normal cholesterol.^3^ Another study found that risk of poor memory was associated with lower high-density lipoprotein cholesterol (HDL-C).^4^ However, results have been inconsistent across studies and the causal relationship has not been rigorously assessed.^5–7^ Blood cholesterol levels have a strong inherited basis. A recent genome-wide association study (GWAS) identified 157 loci associated with blood cholesterol levels, which cumulatively explained over 50% of the inter-individual variation of blood cholesterol levels.^8,9^ Given the large genetic component, genetic predisposition to increased blood cholesterol levels may be used as an instrumental variable to test the inferred causality of blood cholesterol levels on cognitive impairment.

Our current study hypothesized that blood cholesterol levels (HDL-C and TC) play an etiological role in dementia. This hypothesis posits that genetic variants that affect lipid metabolism would influence risk of cognitive impairment through changes in blood cholesterol levels. We applied an inverse□variance weighted Mendelian randomization framework analysis in European and African genetic ancestry samples in a United States based aging study, to investigate the causal nature of lipid dysregulation and cognitive impairment.

## Methods

### Study sample from the Health and Retirement Study

The Health and Retirement Study (HRS), funded by the National Institute on Aging and the Social Security Administration, is a publicly available longitudinal panel cohort study of people over age 50 in the United States. The study includes assessments of economic conditions, health, and other aspects of life surveyed in waves every two years since its inception in 1992.More than 43,000 people have participated in the study to date.^10^ Each wave interviews roughly 20,000 individuals.

Sample selection steps are shown in **Supplementary Figure 1**. We excluded participants who were younger than 50 or over 90 at their cholesterol measurement because the underlying neuropathological mechanisms and risk factors of dementia are considerably different in those age groups.^11^ To minimize misclassification of cognitive status, we also excluded individuals who presented with dementia at the prior wave and with a normal cognition measure in a subsequent wave (n = 29).

### Exposure assessments

HRS began collecting blood-based biomarkers, including HDL-C and TC, from individuals who participated in an enhanced face-to-face interview in 2006 (a randomly selected half of the sample) and 2008 (the other half). Similarly, new cohort members in 2010 were randomly assigned to one of the two groups. The HRS repeats blood sampling for biomarker measures for each group every four years.^12–14^ Special informed consent was obtained for the blood acquisition process.^10^ Cholesterol levels were retrieved from the HRS health sensitive data.^15^ We used the National Health and Nutrition Examination Survey equivalent assay values constructed by the HRS in our analyses.^16^ We selected the first non-missing blood assay results from participants (over the course of four waves with biomarkers 2006 to 2012) to maximize the sample size of non-missing data in biomarkers, genetics, and cognitive assessments across waves (n = 18,700).

We dichotomized blood cholesterol levels based on the 2018 guidelines published in the Journal of the American College of Cardiology:^17^ For HDL-C, normal levels were defined as: ≥40 mg/dL for males or ≥50 mg/dL for females, and at risk (low) levels were defined as: <40 mg/dL for males or <50 mg/dL for females. For TC, normal levels were defined as: <240 mg/dL, and at risk (high) levels were defined as: ≥240 mg/dL. Diverse lipid ratios (e.g. TC/HDL-C, LDL/HDL) have been used as atherogenic indexes for metabolic syndrome and cardiovascular disease. These indices have shown better predictive capacity for the aforementioned diseases than isolated lipid markers.^18^ Hence, we used the TC/HDL-C ratio as a sensitivity analysis in our study.

### Outcome assessments

HRS respondents who could participate in the interview themselves were asked to perform a series of cognition tests (**Table 1**). Their performance on each task was recorded in a continuous scale.^19^ Our main outcome, the Langa-Weir cognitive status, was classified in three levels based on a total score on a 27-point scale (normal: 12-27, cognitive impairment-non dementia (CIND): 7-11, and dementia: 0-6).^20^ Few studies have specifically examined how cholesterol levels influence different cognitive domains, and while the majority have reported that lower HDL levels are tied to worse memory performance, some have reported a lack of association with other cognitive functions.^21^ Thus, we also used summary scores of three separate cognitive domains: word recall for episodic memory, serial 7 subtraction test for mental status, and an adaptation of the Wechsler Adult Intelligence Scale-revised vocabulary in our analyses.^22^

**Table 1.** Cognition domain scores and summary scores in the Health and Retirement Study^a^

|  | Total cognition | Langa-Weir Classification | Domain Analysis |
| --- | --- | --- | --- |
| **Episodic memory** |  |  |  |
| Immediate word recall (0-10) | X | X | X |
| Delayed word recall (0-10) | X | X | X |
| **Mental status** |  |  |  |
| Serial 7 subtraction (0-5) | X | X | X |
| Backward count from 20 (0-2) | X | X | X |
| Date naming (0-4) | X |  |  |
| Object naming (0-2) | X |  |  |
| Naming the president and the vice president of the United States (0-2) | X |  |  |
| **Vocabulary** |  |  |  |
| Vocabulary summary score (0-10) |  |  | X |
| **Total** | 35 | 27 | - |

**Notes:**
[a] All scores were reported in integers, larger score indicates better performance in a certain test.

#### Immediate and delayed recall

The interviewer read a list of 20 nouns (e.g., lake, car, army, etc.) to the respondent, and asked the respondent to recall as many words as possible from the list in any order. After approximately 5 minutes of asking other survey questions (e.g., depression, and cognition items including backwards count, and serial 7’s) the respondent was asked to recall the nouns previously presented as part of the initial recall task.

#### Serial 7’s Test

The interviewer asked the respondent to subtract 7 from 100, and continue subtracting 7 from each subsequent number for a total of five trials. It was up to the respondent to remember the value from the prior subtraction without prompting.

#### Vocabulary

This measure was adapted from the Wechsler Adult Intelligence Scale-revised. Specifically, respondents were asked to define five words from one of two sets: 1) repair, fabric, domestic, remorse, plagiarize, and 2) conceal, enormous, perimeter, compassion, audacious. Respondents are randomly assigned to one set of words in the first wave and the sets are alternated in each wave thereafter.^23^

All cognition variables were measured at the same wave as the corresponding cholesterol measure and retrieved from the HRS cross-wave imputation of cognitive functioning data.^24^

### Covariate assessments

Other covariates used in our analysis included demographic characteristics, behavioral risk factors, and chronic health conditions. Age (years) at was calculated by subtracting the birth year from the cholesterol measurement year. Cholesterol measurement wave was an indicator of individual’s first non-missing cholesterol measure. Sex (male/female), years of education, proxy status (self/proxy-respondent), body mass index (BMI, kilograms/meters^2^), lipid-lowering medication (yes/no), histories of stroke, hypertension, diabetes (yes/no), smoking status (current, former, never), and alcohol consumption (ever drinking yes/no) were self-reported. All covariates were assessed at the cholesterol measurement wave and retrieved from the RAND HRS Longitudinal File.^25^

### Genetic data

HRS began collecting genetic data from respondents in 2006. Respondents provided saliva samples after reading and signing a consent form during an enhanced face-to-face interview. Details of the genotype collection and quality control can be found elsewhere.^26^ Genotyping was conducted by the Center for Inherited Disease Research using the Illumina HumanOmini2.5 BeadChip. Genotype data that passed initial quality control were released and analyzed by the Quality Assurance/Quality Control analysis team at the University of Washington. Raw genetic data on unrelated individuals, both genotyped and imputed to the 1000 Genomes Project, is available from the National Center for Biotechnology Information’s database of genotypes and phenotypes (dbGaP Study Accession: phs000428.v2.p2).

Genetic ancestry in HRS was identified through the union of self-reported race/ethnicity and principal component (PC) analysis on genome-wide single nucleotide polymorphisms (SNP) calculated across all participants plus HapMap controls.^26^ Participants who were self-identified non-Hispanic White and fell within the European ancestry genetic PC cluster were included in the European ancestry sample. Participants who self-identified as non-Hispanic Black and fell within the African ancestry genetic PC cluster were included in the African ancestry sample. The HRS releases ancestry-specific PCs created within each ancestry sample.^27^

We used a polygenic score (PGS) constructed by the HRS as the instrumental variable in our Mendelian randomization analyses. A PGS is a weighted sum of cumulative genetic risk for a trait, which aggregates multiple individual loci across the human genome and weights them by effect sizes from a prior GWAS meta-analysis.^27^ The HDL-C and TC PGSs were created using weights from a 2013 GWAS by the Global Lipid Genetics Consortium.^9^ The general cognition PGS was created using weights from a 2015 GWAS by the Cohorts for Heart and Aging Research in Genomic Epidemiology consortium.^28^ All the PGSs were standardized (mean = 0, standard deviation = 1) within ancestry. We included *APOE*-*ε4* allele carrier status along with the general cognition PGS in our sensitivity analyses as a precision variables to reduce standard errors in the regression models and to capture the large genetic component of dementia attributed to the *APOE*-*ε4* variant. A binary variable of *APOE*-*ε4* allele carrier (having at least one copy of the *ε4* allele, yes/no), was retrieved from the HRS genetic data imputed to the worldwide 1000 Genomes Project reference panel (phase I).^29^ A promoter variant (rs3764261[A]) on the cholesteryl ester transfer protein (*CETP*) gene is consistently related to the metabolism of HDL-C.^34^ We retrieved the number of copies of this SNP from the HRS measured genotype files for sensitivity analyses.

### Statistical analysis

Distributions of baseline characteristics were compared between included and excluded samples, ancestry, and across exposure (cholesterol level) and outcome (cognitive status) groups.χ^2^ test and analysis of variance were used as appropriate to examine the homogeneity across groups. We used linear regressions to examine the associations between cholesterol PGSs and other covariates.

In our main analyses (European ancestry sample), we used multivariable logistic regressions and Wald-type ratio estimators to test the associations and inferred causality between dyslipidemia and cognitive status, using cholesterol PGSs as instrumental variables. Results from the Mendelian randomization and multivariable logistic regression were compared using a test of interaction to evaluate heterogeneity.^30^ Normal cognitive status, normal HDL-C levels, and normal TC levels were treated as reference groups. Our primary models were adjusted for age, sex, years of education, lipid-lowering medication, cholesterol measurement wave, and five ancestry-specific PCs. Additional adjustment included health risk factors that were associated with blood cholesterol levels. We also added *APOE*-*ε4* allele carrier and general cognition PGS in our sensitivity analyses as further potential confounders of the association between lipid profile and dementia risk. We investigated continuous cognitive domain scores as outcomes with linear regression models to understand the associations between cholesterol levels and different cognitive domains.

### Sensitivity analyses

To assess the robustness of our findings, we performed several analyses. Because only the relevance assumption of Mendelian randomization is testable,^31,32^ we performed improvement χ^2^ tests to evaluate the strength of the instrument, with values greater than 10 being taken as evidence for strong instruments.^33^ We checked for pleiotropy to tentatively assess the violation of independence and exclusion restriction assumptions. Hence, we examined the associations between HDL or TC PGS and potential confounding factors using linear regression, as well as the associations between general cognition PGS and HDL PGS or TC PGS, adjusted for five-ancestry-specific PCs. To control for pleiotropy, we replaced the PGS with an indicator variable of variants in the *CETP* SNP (0/1/2 copies of risk allele[A]) as an instrumental variable. As a negative control, we also conducted analyses in subsets of individuals with normal HDL-C or TC levels who were not using any lipid-lowering medication.

We checked for non-linear associations between dyslipidemia and cognitive status by using a smooth function of continuous cholesterol. We examined the association between TC/HDL-C ratio (logarithmic transformed) and cognitive status. Finally, all analyses were additionally conducted in an African ancestry sample.We reported odds ratios (OR) for logistic regression and 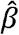 coefficients for linear regression along with their 95% confidence intervals (CIs). Population attributable fractions were also calculated for significant associations. We considered *P*-value <.05 for statistical significance if not specified. All analyses were carried out separately by genetic ancestry and adjusted for a set of five ancestry-specific PCs to adjust for population stratification. A heuristic model and study subsets are shown in **Figure 1**. Analyses were performed in R statistical software (version 3.6.1).^35^ Code to produce all analyses in this manuscript are available online (https://github.com/bakulskilab).

**Figure 1.**
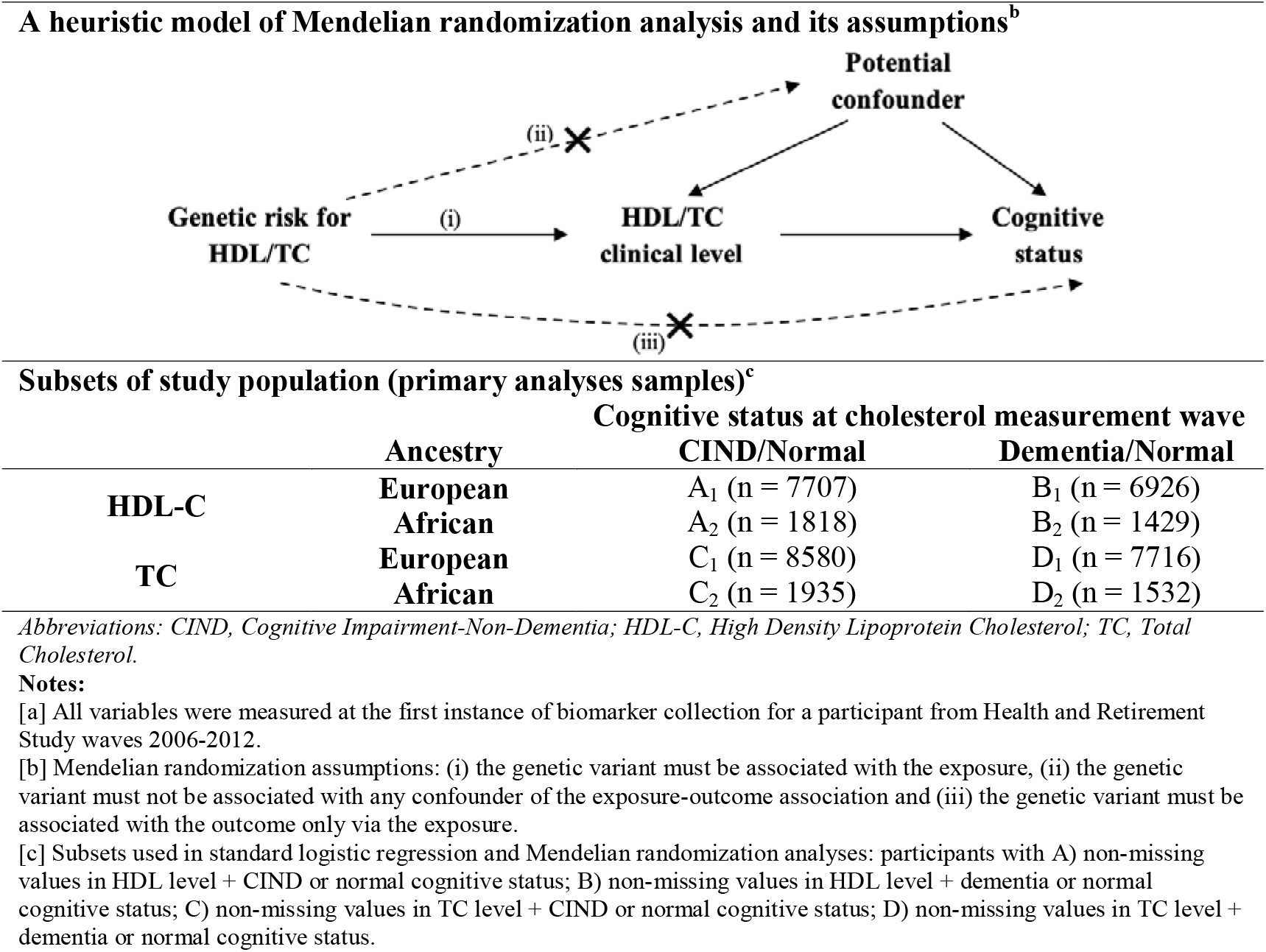
Mendelian randomization analyses structure and subsets of study population, Health and Retirement Study, (n = 10,882)^a^

## Results

There were 10,882 participants included in our analytic sample. Compared to the excluded sample (n = 7,818), our included sample was older, more educated, more likely to have normal TC levels, and had better cognition (**Supplementary Table 1**). Among the included sample, the European and African ancestry groups had similar distributions of HDL-C and TC levels (**Table 2**). Participants in the European ancestry sample were older, more educated, performed better in cognition tests, and less likely to have chronic health conditions, relative to the African ancestry sample.

**Table 2.** Characteristics of sample participants (n = 10,882) stratified by ancestry, in the Health and Retirement Study^a^

|  | Overall<br>n = 10,882 | European<br>n = 8781 | African<br>n = 2101 | Overall<br>P-value<sup>b</sup> |
| --- | --- | --- | --- | --- |
| **Categorical Variables [Count (Frequency)]** |  |  |  |  |
| HDL clinical level<sup>c</sup> |  |  |  | 0.46 |
| Normal | 6711 (68.3%) | 5362 (68.1%) | 1349 (69.0%) |  |
| At risk (low) | 3112 (31.7%) | 2507 (31.9%) | 605 (31.0%) |  |
| TC clinical level<sup>c</sup> |  |  |  | 0.43 |
| Normal | 9170 (84.4%) | 7415 (84.5%) | 1755 (83.8%) |  |
| At risk (high) | 1700 (15.6%) | 1360 (15.5%) | 340 (16.2%) |  |
| Cognitive Status at HDL or TC measurement wave |  |  |  | <0.001* |
| Normal | 8913 (81.9%) | 7537 (85.8%) | 1376 (65.5%) |  |
| Cognitive Impairment-Non Dementia | 1624 (14.9%) | 1059 (12.1%) | 565 (26.9%) |  |
| Dementia | 345 (3.17%) | 185 (2.11%) | 160 (7.62%) |  |
| Sex (Female) | 6423 (59.0%) | 5101 (58.1%) | 1322 (62.9%) | <0.001* |
| Lipid-lowering medication (Yes) | 5430 (49.9%) | 4413 (50.3%) | 1017 (48.4%) | 0.13 |
| Stroke history (Yes) | 824 (7.57%) | 619 (7.05%) | 205 (9.76%) | <0.001* |
| Hypertension history (Yes) | 7150 (65.7%) | 5384 (61.3%) | 1766 (84.1%) | <0.001* |
| Diabetes history (Yes) | 2494 (22.9%) | 1740 (19.8%) | 754 (35.9%) | <0.001* |
| Smoking status |  |  |  | <0.001* |
| Never | 4554 (41.8%) | 3712 (42.3%) | 842 (40.1%) |  |
| Former | 4793 (44.0%) | 3969 (45.2%) | 824 (39.2%) |  |
| Current | 1535 (14.1%) | 1100 (12.5%) | 435 (20.7%) |  |
| Drink status (Ever drinker) | 5922 (54.4%) | 4986 (56.8%) | 936 (44.6%) | <0.001* |
| Proxy status (Self-respondent) | 10882 (100%) | 8781 (100%) | 2101 (100%) | . |
| APOE-ε4 allele carrier (Yes) | 2605 (27.8%) | 2003 (25.8%) | 602 (37.9%) | <0.001* |
| Cholesterol measure wave |  |  |  | <0.001* |
| Wave 2006 | 3893 (35.8%) | 3394 (38.7%) | 499 (23.8%) |  |
| Wave 2008 | 3922 (36.0%) | 3341 (38.0%) | 581 (27.7%) |  |
| Wave 2010 | 1582 (14.5%) | 1063 (12.1%) | 519 (24.7%) |  |
| Wave 2012 | 1485 (13.6%) | 983 (11.2%) | 502 (23.9%) |  |
| **Continuous Variables [Mean (SD)]** |  |  |  |  |
| Blood HDL (mg/dL) | 54.4 (16.1) | 54.2 (16.1) | 55.0 (16.0) | 0.06 |
| Blood TC (mg/dL) | 197 (42.2) | 197 (42.2) | 197 (42.3) | 0.67 |
| Age at HDL or TC measurement wave (yrs) | 67.6 (10.1) | 68.4 (10.1) | 64.1 (9.34) | <0.001* |
| Years of education | 13.0 (2.65) | 13.2 (2.54) | 12.2 (2.91) | <0.001* |
| Body Mass Index (kg/m<sup>2</sup>) | 28.9 (6.16) | 28.4 (5.84) | 30.8 (7.02) | <0.001* |
| Immediate word recall | 5.50 (1.58) | 5.61 (1.57) | 5.06 (1.52) | <0.001* |
| Delayed word recall | 4.36 (1.92) | 4.53 (1.91) | 3.65 (1.83) | <0.001* |
| Serial 7 subtraction | 3.57 (1.64) | 3.82 (1.50) | 2.54 (1.80) | <0.001* |
| Backward count from 20 | 1.89 (0.46) | 1.92 (0.40) | 1.77 (0.64) | <0.001* |
| Vocabulary sum score | 5.56 (2.05) | 5.86 (1.89) | 4.34 (2.19) | <0.001* |
| Total cognition | 22.1 (4.85) | 22.6 (4.57) | 19.3 (5.18) | <0.001* |

Abbreviations: APOE, Apolipoprotein E; HDL, High Density Lipoprotein Cholesterol; SD: Standard Deviation; TC, Total Cholesterol.
**Notes:**
[a] All variables were measured at the first instance of biomarker collection for a participant from Health and Retirement Study waves 2006-2012. All the statistics including count, frequency, mean, SD, and *P* value were calculated based on non-missing data for each variable.
[b] The overall *P* value was calculated from chi-square test or analysis of variance for categorical or continuous variables as appropriate, interpreted as differences between groups. \* indicates a significance at a *P* value of 0.05.
[c] At risk low HDL: <40 mg/dL for male and <50 mg/dL for female; At risk high TC: ≥240 mg/dL.

In the European ancestry sample (n = 8,781), health status including BMI, smoking, alcohol consumption, histories of stroke, hypertension, and diabetes were associated with both outcome and exposures. Thus in our sensitivity models, we additionally adjusted for these variables. Cognitive status was positively associated with the *APOE*-*ε4* allele carrier status and general cognition PGS (**Table 3)**.

**Table 3.** Bivariate characteristics stratified by cognitive status or cholesterol clinical level, in the Health and Retirement Study, European ancestry sample (n = 8781)^a^

|  | Cognitive Status |  |  |  |  | High Density Lipoprotein Cholesterol (HDL) |  |  |  | Total Cholesterol (TC) |  |  |  |
| --- | --- | --- | --- | --- | --- | --- | --- | --- | --- | --- | --- | --- | --- |
|  | Overall<br>n = 8781 | Normal<br>n = 7537 | CIND<br>n = 1059 | Dementia<br>n = 185 | P-value<sup>b</sup> | Overall<br>n = 7869 | Normal<br>n = 5362 | At risk (low)<br>n = 2507 | P-value<sup>b</sup> | Overall<br>n = 8775 | Normal<br>n = 7415 | At risk (high)<br>n = 1360 | P-value<sup>b</sup> |
| **Categorical Variables [Count (Frequency)]** |  |  |  |  |  |  |  |  |  |  |  |  |  |
| HDL clinical level<sup>c</sup> |  |  |  |  | <0.001* |  |  | - |  |  |  |  | <0.001* |
| Normal | 5362 (68.1%) | 4668 (69.0%) | 590 (62.6%) | 104 (64.2%) |  |  |  |  |  | 5358 (68.1%) | 4288 (65.2%) | 1070 (83.1%) |  |
| At risk (low) | 2507 (31.9%) | 2096 (31.0%) | 353 (37.4%) | 58 (35.8%) |  |  |  |  |  | 2505 (31.9%) | 2287 (34.8%) | 218 (16.9%) |  |
| TC clinical level<sup>c</sup> |  |  |  |  | 0.12 |  |  |  | <0.001* |  |  | - |  |
| Normal | 7415 (84.5%) | 6340 (84.2%) | 913 (86.2%) | 162 (87.6%) |  | 6575 (83.6%) | 4288 (80.0%) | 2287 (91.3%) |  |  |  |  |  |
| At risk (high) | 1360 (15.5%) | 1191 (15.8%) | 146 (13.8%) | 23 (12.4%) |  | 1288 (16.4%) | 1070 (20.0%) | 218 (8.70%) |  |  |  |  |  |
| Sex (Female) | 5101 (58.1%) | 4433 (58.8%) | 570 (53.8%) | 98 (53.0%) | 0.003* | 4599 (58.4%) | 3070 (57.3%) | 1529 (61.0%) | 0.002* | 5099 (58.1%) | 4083 (55.1%) | 1016 (74.7%) | <0.001* |
| Lipid-lowering medication (Yes) | 4413 (50.3%) | 3735 (49.6%) | 588 (55.5%) | 90 (48.6%) | 0.001* | 3958 (50.3%) | 2592 (48.3%) | 1366 (54.5%) | <0.001* | 4412 (50.3%) | 4009 (54.1%) | 403 (29.6%) | <0.001* |
| Stroke history (Yes) | 619 (7.05%) | 414 (5.49%) | 163 (15.4%) | 42 (22.7%) | <0.001* | 553 (7.03%) | 326 (6.08%) | 227 (9.05%) | <0.001* | 619 (7.05%) | 549 (7.40%) | 70 (5.15%) | 0.003* |
| Hypertension history (Yes) | 5384 (61.3%) | 4547 (60.3%) | 721 (68.1%) | 116 (62.7%) | <0.001* | 4814 (61.2%) | 3133 (58.4%) | 1681 (67.1%) | <0.001* | 5380 (61.3%) | 4623 (62.3%) | 757 (55.7%) | <0.001* |
| Diabetes history (Yes) | 1740 (19.8%) | 1428 (18.9%) | 264 (24.9%) | 48 (25.9%) | <0.001* | 1533 (19.5%) | 839 (15.6%) | 694 (27.7%) | <0.001* | 1739 (19.8%) | 1591 (21.5%) | 148 (10.9%) | <0.001* |
| Smoking status |  |  |  |  | 0.001* |  |  |  | <0.001* |  |  |  | <0.001* |
| Never | 3712 (42.3%) | 3243 (43.0%) | 386 (36.4%) | 83 (44.9%) |  | 3306 (42.0%) | 2285 (42.6%) | 1021 (40.7%) |  | 3710 (42.3%) | 3091 (41.7%) | 619 (45.5%) |  |
| Former | 3969 (45.2%) | 3361 (44.6%) | 524 (49.5%) | 84 (45.4%) |  | 3550 (45.1%) | 2444 (45.6%) | 1106 (44.1%) |  | 3968 (45.2%) | 3427 (46.2%) | 541 (39.8%) |  |
| Current | 1100 (12.5%) | 933 (12.4%) | 149 (14.1%) | 18 (9.73%) |  | 1013 (12.9%) | 633 (11.8%) | 380 (15.2%) |  | 1097 (12.5%) | 897 (12.1%) | 200 (14.7%) |  |
| Drink status (Ever drinker) | 4986 (56.8%) | 4492 (59.6%) | 438 (41.4%) | 56 (30.3%) | <0.001* | 4479 (56.9%) | 3262 (60.8%) | 1217 (48.5%) | <0.001* | 4982 (56.8%) | 4189 (56.5%) | 793 (58.3%) | 0.23 |
| APOE-ε4 allele carrier (Yes) | 2003 (25.8%) | 1659 (24.9%) | 274 (28.8%) | 70 (41.2%) | <0.001* | 1787 (25.9%) | 1206 (25.8%) | 581 (26.2%) | 0.74 | 2001 (25.7%) | 1656 (25.3%) | 345 (28.1%) | 0.04* |
| First cholesterol measure wave |  |  |  |  | 0.91 |  |  |  | 0.318 |  |  |  | 0.02* |
| Wave 2006 | 3394 (38.7%) | 2910 (38.6%) | 412 (38.9%) | 72 (38.9%) |  | 2758 (35.0%) | 1857 (34.6%) | 901 (35.9%) |  | 3394 (38.7%) | 2858 (38.5%) | 536 (39.4%) |  |
| Wave 2008 | 3341 (38.0%) | 2853 (37.9%) | 417 (39.4%) | 71 (38.4%) |  | 3086 (39.2%) | 2094 (39.1%) | 992 (39.6%) |  | 3341 (38.1%) | 2792 (37.7%) | 549 (40.4%) |  |
| Wave 2010 | 1063 (12.1%) | 924 (12.3%) | 117 (11.0%) | 22 (11.9%) |  | 1051 (13.4%) | 726 (13.5%) | 325 (13.0%) |  | 1058 (12.1%) | 905 (12.2%) | 153 (11.2%) |  |
| Wave 2012 | 983 (11.2%) | 850 (11.3%) | 113 (10.7%) | 20 (10.8%) |  | 974 (12.4%) | 685 (12.8%) | 289 (11.5%) |  | 982 (11.2%) | 860 (11.6%) | 122 (8.97%) |  |
| **Continuous Variables [Mean (SD)]** |  |  |  |  |  |  |  |  |  |  |  |  |  |
| Age at measurement (yrs) | 68.4 (10.1) | 67.4 (9.72) | 74.5 (9.99) | 76.6 (9.26) | <0.001* | 68.1 (10.1) | 67.9 (10.2) | 68.7 (10.0) | 0.001* | 68.4 (10.1) | 68.8 (10.1) | 66.4 (9.65) | <0.001* |
| Years of education | 13.2 (2.52) | 13.5 (2.34) | 11.7 (2.76) | 10.6 (3.32) | <0.001* | 13.2 (2.51) | 13.4 (2.52) | 12.9 (2.46) | <0.001* | 13.2 (2.52) | 13.2 (2.53) | 13.3 (2.42) | 0.50 |
| HDL polygenic risk score | -0.01 (1.00) | -0.01 (1.00) | 0.01 (0.98) | -0.16 (1.06) | 0.11 | -0.02 (1.00) | 0.06 (0.99) | -0.20 (1.01) | <0.001* | -0.01 (1.00) | -0.03 (1.01) | 0.08 (0.97) | <0.001* |
| TC polygenic risk score | 0.00 (1.00) | 0.00 (1.00) | -0.02 (0.99) | -0.08 (1.04) | 0.42 | -0.01 (0.99) | 0.02 (0.99) | -0.06 (0.98) | <0.001* | 0.00 (1.00) | -0.02 (1.00) | 0.09 (0.98) | <0.001* |
| General cognition polygenic risk score | -0.01 (1.00) | 0.00 (1.00) | -0.11 (0.98) | 0.01 (0.92) | 0.002* | -0.01 (1.00) | 0.00 (1.01) | -0.05 (0.97) | 0.04* | -0.01 (1.00) | -0.01 (1.00) | -0.03 (1.00) | 0.36 |
| BMI (kg/m<sup>2</sup>) | 28.4 (5.84) | 28.6 (5.82) | 27.2 (5.82) | 26.8 (5.84) | <0.001* | 28.4 (5.88) | 27.8 (5.61) | 29.9 (6.17) | <0.001* | 28.4 (5.84) | 28.5 (5.87) | 27.9 (5.68) | 0.001* |
| Immediate word recall | 5.61 (1.57) | 5.97 (1.32) | 3.69 (1.05) | 2.03 (1.20) | <0.001* | 5.62 (1.56) | 5.69 (1.57) | 5.48 (1.55) | <0.001* | 5.61 (1.57) | 5.58 (1.57) | 5.76 (1.55) | <0.001* |
| Delayed word recall | 4.54 (1.90) | 4.99 (1.57) | 2.02 (1.35) | 0.53 (0.77) | <0.001* | 4.55 (1.90) | 4.63 (1.90) | 4.39 (1.90) | <0.001* | 4.54 (1.90) | 4.49 (1.90) | 4.78 (1.90) | <0.001* |
| Serial 7 subtraction | 3.82 (1.50) | 4.14 (1.21) | 2.08 (1.63) | 0.64 (0.97) | <0.001* | 3.81 (1.51) | 3.86 (1.47) | 3.72 (1.58) | <0.001* | 3.82 (1.50) | 3.83 (1.49) | 3.73 (1.56) | 0.02* |
| Backward count from 20 | 1.92 (0.39) | 1.95 (0.31) | 1.79 (0.61) | 1.32 (0.95) | <0.001* | 1.92 (0.39) | 1.92 (0.39) | 1.92 (0.39) | 0.87 | 1.92 (0.39) | 1.92 (0.40) | 1.93 (0.37) | 0.44 |
| Vocabulary sum score | 5.87 (1.89) | 6.09 (1.80) | 4.98 (1.87) | 3.81 (2.10) | <0.001* | 5.87 (1.91) | 5.89 (1.89) | 5.81 (1.94) | 0.32 | 5.86 (1.89) | 5.86 (1.89) | 5.89 (1.90) | 0.79 |
| Total cognition | 22.6 (4.54) | 24.2 (3.05) | 16.6 (1.92) | 10.0 (3.42) | <0.001* | 22.7 (4.54) | 22.9 (4.51) | 22.3 (4.58) | <0.001* | 22.6 (4.54) | 22.6 (4.51) | 23.0 (4.72) | 0.01* |

Abbreviations: APOE, Apolipoprotein E; CIND, Cognitive Impairment Non-Dementia; HDL, High Density Lipoprotein Cholesterol; SD, Standard Deviation; TC, Total Cholesterol.
**Notes:**
[a] All variables were measured at the first instance of biomarker collection for a participant from Health and Retirement Study waves 2006-2012. All the statistics including count, frequency, mean, SD, and P value were calculated based on non-missing data for each variable.
[b] The overall P value was calculated from chi-square test or analysis of variance for categorical or continuous variables as appropriate, interpreted as differences between groups. \* indicates a significance at a P value of 0.05.
[c] At risk low HDL: <40 mg/dL for male and <50 mg/dL for female; At risk high TC: ≥240 mg/dL.

### Associations with cumulative genetic risk of dyslipidemia

In the European ancestry sample, cholesterol PGSs were highly associated with their corresponding blood cholesterol measures. After adjusting for age, sex, years of education, lipid-lowering medication, cholesterol measurement wave, and five ancestry-specific PCs, a one standard deviation increase in HDL PGS was associated with 0.78 (95%CI: 0.74, 0.82) lower odds of at risk relative to normal HDL-C level; a one standard deviation unit increase in TC PGS was associated with 1.21 (95%CI: 1.13, 1.29) times odds of at risk relative to normal TC level.Improvement χ^2^ test statistics confirmed both cholesterol PGSs as valid instruments for blood cholesterol levels in our sample (HDL PGS: χ^2^=98.7; TC PGS: χ^2^=32.6) (**Table 4**).

**Table 4.** Associations between polygenic risk score for cholesterol (HDL-C and TC) and blood cholesterol levels, in the Health and Retirement Study, European ancestry sample (n = 8781)^a^

|  | CIND & Normal |  |  | Dementia & Normal |  |  | Overall sample |  |  |
| --- | --- | --- | --- | --- | --- | --- | --- | --- | --- |
|  | N | OR | (95 CI%) | N | OR | (95 CI%) | N | OR | (95 CI%) |
| **High Density Lipoprotein Cholesterol (HDL-C)** |  |  |  |  |  |  |  |  |  |
| Crude | 7707 | 0.77 | (0.73, 0.81) | 6926 | 0.75 | (0.71, 0.79) | 7997 | 0.77 | (0.73, 0.81) |
| Adjusted<sup>b</sup> | 7707 | 0.78 | (0.74, 0.82) | 6926 | 0.77 | (0.73, 0.81) | 7997 | 0.78 | (0.74, 0.82) |
| **Improvement $\chi^2$<sup>c</sup>** |  |  | 97.5 |  |  | 99.8 |  |  | 98.7 |
| **Total Cholesterol (TC)** |  |  |  |  |  |  |  |  |  |
| Crude | 8590 | 1.12 | (1.05, 1.18) | 7716 | 1.10 | (1.03, 1.17) | 8921 | 1.11 | (1.05, 1.18) |
| Adjusted | 8590 | 1.21 | (1.13, 1.30) | 7716 | 1.20 | (1.12, 1.28) | 8921 | 1.21 | (1.13, 1.29) |
| **Improvement $\chi^2$** |  |  | 32.6 |  |  | 25.5 |  |  | 32.6 |

Abbreviations: CI, Confidence Interval; CIND, Cognitive Impairment-Non Dementia; OR, Odds Ratio
**Notes:**
[a] All variables were measured at the first instance of biomarker collection for a participant from Health and Retirement Study waves 2006-2012. All the values were based on results from multivariable logistic regression analyses in each sample, in which "normal cholesterol level" was used as the reference group.
[b] Adjusted for age, sex, years of education, lipid-lowering medication, cholesterol measurement wave, and five ancestry-specific principal components.
[c] Calculated by $2 \times (\log \text{likelihood of full model} - \log \text{likelihood of reduced model})$ . Statistics larger than 10 indicates a valid instrument in convention.

In the European ancestry sample, HDL PGS was not associated with CIND but was associated with dementia (**Table 5**). After adjusting for age, sex, years of education, lipid-lowering medication, cholesterol measurement wave, and five ancestry-specific PCs, a one standard deviation increase in HDL PGS was associated with 0.81 (95%CI: 0.69, 0.96) lower odds of dementia relative to normal cognition. This association attenuated after additional adjustments of health status (**Table 5**). TC PGS was not directly or indirectly associated with the odds of CIND or dementia. No association was observed between HDL or TC PGS and any cognitive domain score (**Supplementary Table 2**).

**Table 5.** Associations between polygenic risk score for cholesterol and cognitive status, in the Health and Retirement Study, European ancestry sample (n = 8781)^a^

|  | High Density Lipoprotein Cholesterol (HDL) |  |  |  |  |  | Total Cholesterol (TC) |  |  |  |  |  |
| --- | --- | --- | --- | --- | --- | --- | --- | --- | --- | --- | --- | --- |
|  | CIND vs. Normal |  |  | Dementia vs. Normal |  |  | CIND vs. Normal |  |  | Dementia vs. Normal |  |  |
|  | N | OR | (95 CI%) | N | OR | (95 CI%) | N | OR | (95 CI%) | N | OR | (95 CI%) |
| **Total effect of cholesterol PGS** |  |  |  |  |  |  |  |  |  |  |  |  |
| Crude | 7707 | 1.03 | (0.96, 1.10) | 6926 | 0.83 | (0.71, 0.97) | 8590 | 0.98 | (0.92, 1.04) | 7716 | 0.92 | (0.79, 1.06) |
| Adjusted (demographic)<sup>b</sup> | 7707 | 1.02 | (0.95, 1.10) | 6926 | 0.82 | (0.69, 0.96) | 8590 | 0.97 | (0.90, 1.05) | 7716 | 0.90 | (0.76, 1.07) |
| Adjusted (health status)<sup>c</sup> | 7707 | 1.03 | (0.96, 1.11) | 6926 | 0.83 | (0.70, 0.98) | 8590 | 0.97 | (0.90, 1.05) | 7716 | 0.90 | (0.76, 1.07) |
| **Direct effect of cholesterol PGS (adjusting for cholesterol level)** |  |  |  |  |  |  |  |  |  |  |  |  |
| Crude | 7707 | 1.05 | (0.98, 1.12) | 6926 | 0.84 | (0.72, 0.98) | 8590 | 0.98 | (0.92, 1.05) | 7716 | 0.92 | (0.79, 1.06) |
| Adjusted (demographic) | 7707 | 1.03 | (0.96, 1.11) | 6926 | 0.81 | (0.69, 0.96) | 8590 | 0.97 | (0.90, 1.04) | 7716 | 0.90 | (0.76, 1.07) |
| Adjusted (health status) | 7707 | 1.04 | (0.97, 1.12) | 6926 | 0.83 | (0.70, 0.98) | 8590 | 0.97 | (0.90, 1.05) | 7716 | 0.91 | (0.76, 1.07) |

Abbreviations: APOE, Apolipoprotein E; CI, Confidence Interval; CIND, Cognitive Impairment-Non Dementia; OR, Odds Ratio
**Notes:**
[a] All variables were measured at the first instance of biomarker collection for a participant from Health and Retirement Study waves 2006-2012. All the values were based on results from multivariable logistic regression analyses in each sample, in which "normal cognitive status" and "normal cholesterol level" were used as the reference groups.
[b] Adjusted for age, sex, years of education, lipid-lowering medication, cholesterol measurement wave, and five ancestry-specific principal components.
[c] Adjusted for ever drink alcohol, history of stroke, hypertension, diabetes, and BMI in addition to variables in [b].

**Figure 2** presents the associations of HDL or TC PGSs with baseline characteristics in the European ancestry sample. Both cholesterol PGSs were associated with lipid-lowering medication usage and alcohol consumption. In addition, HDL PGS was also associated with history of hypertension, diabetes, smoking status, BMI, and years of education.

**Figure 2.**
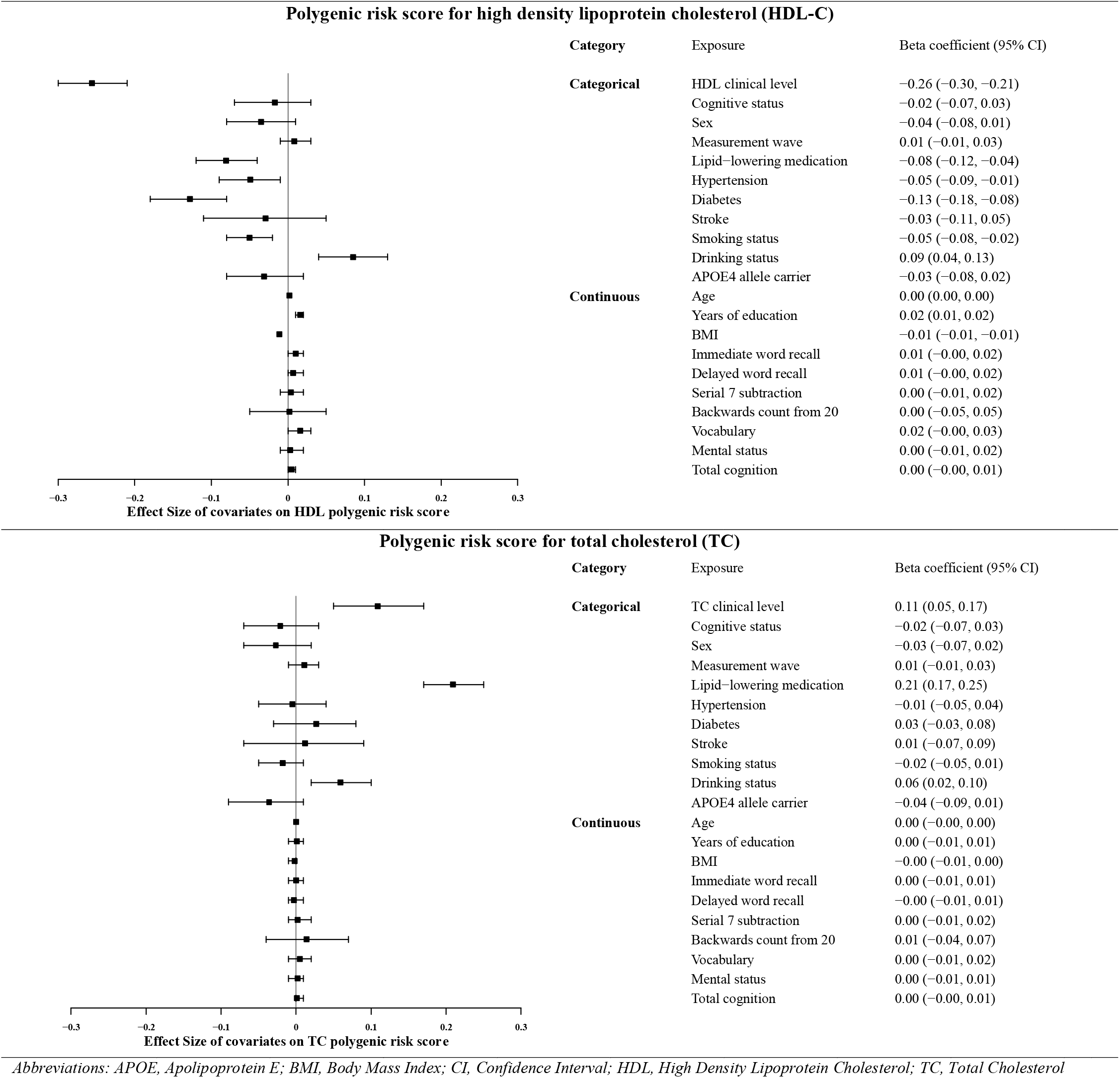
Associations between cholesterol polygenic risk score and factors potentially confounding the relation between cholesterol and cognitive status, in the Health and Retirement Study, European ancestry sample (n = 8781)

### Associations between HDL-C levels and cognition

Multivariable logistic regression results showed at risk (low) HDL-C levels were associated with increased odds of CIND (**Table 6**). According to population attributable fraction results, 5% of the CIND cases were attributed to at risk (low) HDL-C levels. In the primary adjusted model, those with at risk (low) HDL-C levels had 1.20 (95%CI: 1.03, 1.40) times higher odds of CIND relative to normal cognition. This association attenuated to the null after additional adjustments for health status. No associations were observed between HDL-C levels and dementia.

**Table 6.** Associations between blood cholesterol levels and cognitive status, in the Health and Retirement Study, European ancestry sample (n = 8781)^a^

|  | High Density Lipoprotein Cholesterol (HDL) |  |  |  |  |  | Total Cholesterol (TC) |  |  |  |  |  |
| --- | --- | --- | --- | --- | --- | --- | --- | --- | --- | --- | --- | --- |
|  | N | Logistic Regression |  | Wald-type/ratio |  | P for heterogeneity<sup>b</sup> | N | Logistic Regression |  | Wald-type/ratio |  | P for heterogeneity |
|  |  | OR | (95 CI%) | OR | (95 CI%) |  |  | OR | (95 CI%) |  |  |  |
| *CIND vs. Normal* |  |  |  |  |  |  |  |  |  |  |  |  |
| Crude | 7707 | 1.33 | (1.16, 1.53) | 0.89 | (0.69, 1.16) | 0.01* | 8590 | 0.85 | (0.70, 1.02) | 0.82 | (0.45, 1.48) | 0.91 |
| Adjusted (demographic)<sup>c</sup> | 7707 | 1.20 | (1.03, 1.40) | 0.91 | (0.68, 1.23) | 0.11 | 8590 | 1.10 | (0.90, 1.34) | 0.86 | (0.58, 1.26) | 0.27 |
| Adjusted (health status)<sup>d</sup> | 7707 | 1.15 | (0.99, 1.35) | 0.88 | (0.65, 1.18) | 0.12 | 8590 | 1.13 | (0.91, 1.38) | 0.87 | (0.59, 1.28) | 0.17 |
| *Dementia vs. Normal* |  |  |  |  |  |  |  |  |  |  |  |  |
| Crude | 6926 | 1.24 | (0.89, 1.71) | 1.94 | (1.13, 3.34) | 0.16 | 7716 | 0.76 | (0.47, 1.15) | 0.40 | (0.08, 1.88) | 0.44 |
| Adjusted (demographic) | 6926 | 1.02 | (0.72, 1.44) | 2.15 | (1.16, 3.99) | 0.04* | 7716 | 0.93 | (0.57, 1.47) | 0.56 | (0.22, 1.43) | 0.34 |
| Adjusted (health status) | 6926 | 0.97 | (0.67, 1.39) | 2.02 | (1.08, 3.77) | 0.047* | 7716 | 0.91 | (0.55, 1.44) | 0.57 | (0.22, 1.47) | 0.39 |

Abbreviations: APOE, Apolipoprotein E; BMI, Body Mass Index; CI: Confidence Interval; CIND: Cognitive Impairment-Non Dementia; OR: Odds Ratio
**Notes:**
[a] All variables were measured at the first instance of biomarker collection for a participant from Health and Retirement Study waves 2006-2012. All values were based on results from multivariable logistic regression or Mendelian randomization analyses in each sample, in which "normal cognitive status" and "normal cholesterol level" were used as reference groups.
[b] P value represents the statistical significance of the test of heterogeneity between logistic regression and Mendelian randomization. \* indicates a significance at a P value of 0.05.
[c] Adjusted for age, sex, years of education, lipid-lowering medication, cholesterol measurement wave, and five ancestry-specific principal components.
[d] Adjusted for ever drink alcohol, history of stroke, hypertension, diabetes, and BMI in addition to variables in [c].

Mendelian randomization analyses showed no evidence of inferred causality between HDL-C levels and CIND (OR = 0.92, 95%CI: 0.69, 1.23; **Table 6**), despite significant logistic associations observed between HDL-C levels and CIND. However, an inferred causal relationship was observed between HDL-C levels and dementia. According to the Wald-type/ratio results, using HDL PGS as an instrumental variable, individuals with at risk (low) HDL-C levels had 2.15 (95%CI: 1.16, 3.99) times higher odds of dementia relative to normal cognition. This Mendelian randomization result was significantly different from the multivariable logistic regression result (*P* for heterogeneity = 0.04).

For specific cognitive domains, at risk (low) HDL-C levels were associated with worse performance in word recall tests (episodic memory) only in multivariable regression (**Table 7**). No association was observed between HDL-C levels and mental status or vocabulary scores. None of the associations between HDL-C levels and cognitive domain score could be considered causal through Mendelian randomization analyses.

**Table 7.** Associations between blood cholesterol levels and cognitive domain scores, in the Health and Retirement Study, European ancestry sample (n = 8781)^a^

|  |  | High Density Lipoprotein Cholesterol (HDL-C) |  |  |  |  |  | Total Cholesterol (TC) |  |  |  |  |  |
| --- | --- | --- | --- | --- | --- | --- | --- | --- | --- | --- | --- | --- | --- |
|  |  | Linear Regression |  |  | Wald-type/ratio |  | P for heterogeneity<sup>b</sup> | Linear Regression |  |  | Wald-type/ratio |  | P for heterogeneity |
|  |  | N | β coefficient (95% CI) |  | β coefficient (95% CI) |  |  | N | β coefficient (95% CI) |  | β coefficient (95% CI) |  |  |
| Total cognition score (0-35) | Crude | 5270 | -0.61 | (-0.87, -0.35) | -0.38 | (-0.82, 0.06) | 0.19 | 5924 | 0.45 | (0.12, 0.79) | 0.23 | (-0.84, 1.29) | 0.75 |
|  | Adjusted<sup>c</sup> | 5270 | -0.26 | (-0.49, -0.03) | -0.26 | (-0.67, 0.14) | 0.18 | 5924 | -0.20 | (-0.50, 0.10) | 0.20 | (-0.59, 1.00) | 0.52 |
| Episodic memory |  |  |  |  |  |  |  |  |  |  |  |  |  |
| Immediate word recall | Crude | 7869 | -0.21 | (-0.28, -0.13) | -0.10 | (-0.23, 0.02) | 0.26 | 8775 | 0.18 | (0.09, 0.27) | 0.04 | (-0.27, 0.35) | 0.93 |
|  | Adjusted | 7869 | -0.10 | (-0.16, -0.03) | -0.10 | (-0.21, 0.02) | 0.11 | 8775 | -0.08 | (-0.16, 0.00) | 0.05 | (-0.18, 0.28) | 0.57 |
| Delayed word recall | Crude | 7869 | -0.25 | (-0.34, -0.16) | -0.11 | (-0.26, 0.04) | 0.49 | 8775 | 0.29 | (0.18, 0.40) | -0.05 | (-0.42, 0.32) | 0.63 |
|  | Adjusted | 7869 | -0.13 | (-0.21, -0.05) | -0.13 | (-0.27, 0.01) | 0.17 | 8775 | -0.01 | (-0.11, 0.09) | 0.06 | (-0.22, 0.34) | 0.64 |
| Mental status |  |  |  |  |  |  |  |  |  |  |  |  |  |
| Serial 7 subtraction | Crude | 7869 | -0.13 | (-0.20, -0.06) | -0.05 | (-0.17, 0.07) | 0.68 | 8775 | -0.11 | (-0.19, -0.02) | 0.08 | (-0.21, 0.38) | 0.53 |
|  | Adjusted | 7869 | -0.01 | (-0.08, 0.05) | 0.03 | (-0.09, 0.15) | 0.87 | 8775 | -0.09 | (-0.18, -0.01) | 0.06 | (-0.17, 0.29) | 0.66 |
| Backward count from 20 | Crude | 7869 | 0.00 | (-0.02, 0.02) | 0.00 | (-0.04, 0.03) | 0.76 | 8775 | 0.01 | (-0.01, 0.03) | 0.03 | (-0.05, 0.10) | 0.52 |
|  | Adjusted | 7869 | 0.01 | (-0.01, 0.03) | 0.00 | (-0.04, 0.03) | 0.70 | 8775 | 0.00 | (-0.02, 0.02) | 0.03 | (-0.03, 0.09) | 0.46 |
| Vocabulary |  |  |  |  |  |  |  |  |  |  |  |  |  |
| Vocabulary summary score | Crude | 2714 | -0.08 | (-0.23, 0.07) | -0.20 | (-0.45, 0.05) | 0.08 | 3166 | 0.03 | (-0.16, 0.22) | 0.09 | (-0.52, 0.70) | 0.78 |
|  | Adjusted | 2714 | 0.07 | (-0.07, 0.21) | -0.08 | (-0.31, 0.16) | 0.28 | 3166 | -0.09 | (-0.27, 0.09) | -0.07 | (-0.54, 0.40) | 0.96 |

Abbreviations: BMI, Body Mass Index; CI: Confidence Interval; CIND: Cognitive Impairment-Non Dementia; OR: Odds Ratio
**Notes:**
[a] All variables were measured at the first instance of biomarker collection for a participant from Health and Retirement Study waves 2006-2012. All values were based on results from linear regression and Mendelian randomization analyses in each sample.
[b] P value represents the statistical significance of the test of heterogeneity between linear regression and Mendelian randomization. \* indicates a significance at a P value of 0.05.
[c] Adjusted for age, sex, years of education, lipid-lowering medication, cholesterol measurement wave, and five ancestry-specific principal components.

### Associations between TC levels and cognition

Neither significant logistic regression associations nor causal inference were observed between TC levels and cognitive status in either multivariable logistic regression or Mendelian randomization analyses. Results did not differ between the logistic regression and Mendelian randomization estimates (*P* for heterogeneity: CIND/normal = 0.27; dementia/normal = 0.34) **(Table 6)**. For specific cognitive domains, no association or causal relationship was observed between TC levels and any cognitive domain score (**Table 7**).

### Sensitivity analyses

In the European ancestry sample, after adjusting for five ancestry-specific PCs, general cognition PGS was not associated with HDL PGS (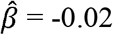, *P* = 0.08), but was associated with TC PGS (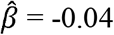, *P* = 0.005). Multivariable logistic regression and Mendelian randomization results were similar when we further adjusted for dementia genetic variables (*APOE*-*ε4* allele carrier status and general cognition PGS).

Given that HDL PGS was associated with multiple baseline characteristics (**Figure 2**), there may be pleiotropic affects between cognition and blood HDL-C levels. To control for this potential pleiotropy, we used rs3764261 as a valid instrumental variable, but no causal relationship was found as above (**Supplementary Table 3**). In the subsample of participants with normal HDL-C and TC levels and not using any lipid-lowering medication, neither HDL PGS nor TC PGS was associated with cognitive status, indicating the independence and exclusion restriction assumptions are likely to hold in our analysis (**Supplementary Table 4**). No association was observed between the log-transformed TC/HDL-C ratio on cognitive status (**Supplementary Table 5**).

**Supplementary Figure 2** shows non-linear associations between continuous cholesterol and cognitive status, adjusting for age, sex, years of education, lipid-lowering medication, cholesterol measurement wave, and five ancestry-specific PCs. The probability of predicted CIND and dementia both decreased with increased HDL-C, which were consistent with our previous findings that low HDL-C level was associated with cognitive impairment. For TC, we observed an expected non-linear association with CIND and dementia: before the clinical cut point of 240 mg/dL, the probability of predicted CIND and dementia both decreases with the increase of TC concentration; while the probability increased after the cut point.

Among the African ancestry sample (n = 2,101), HDL and TC PGSs were also associated with blood HDL-C and TC levels respectively; while improvement χ^2^ test statistics indicated neither cholesterol PGSs were valid instruments (HDL: 4.96 < 10; TC: 9.09 < 10), so no further Mendelian randomization analyses were performed (**Supplementary Table 6a**). In multivariable logistic regression, no associations were observed between HDL or TC polygenic score and CIND or dementia (**Supplementary Table 6b, Model 1**), or between HDL-C or TC level and cognitive status (**Supplementary Table 6b, Model 2**).

## Discussion

The present study was conducted among a sample of older adults from the Health and Retirement Study participating in the 2006 to 2012 waves with biomarker and cognition data. To our knowledge, this is a novel study using a Mendelian randomization framework to investigate the inferred causal nature of dyslipidemia on cognitive status and separate cognitive domains. In the European ancestry sample, after adjusting for age, sex, years of education, lipid-lowing medication, cholesterol measurement wave, and five ancestry-specific PCs, at risk (low) HDL-C levels were associated with higher odds of CIND, lower episodic memory scores, and lower mental status scores. Using inverse□variance weighted Mendelian randomization methods, at risk (low) HDL-C level was inferred to be causally associated with dementia (OR = 2.25, 95%CI: 1.23, 4.09). No associations or causal inferences were observed between TC levels and cognitive status. These findings represent important new information supporting HDL-C management as a public health approach to prevent dementia.

Our observed associations between at risk (low) HDL-C levels and cognitive impairment in the European ancestry sample are consistent with previous studies.^36,37^ HDL-C is involved in the deposition and clearance of beta-amyloid – a determining factor for endothelial inflammation and subsequent neurodegeneration in the brain.^38^ Animal models show a protective effect of HDL-C against memory deficits, neuroinflammation, and cerebral amyloid angiopathy.^39^ Several longitudinal studies also found a potential protective effect of midlife HDL-C on future cognitive decline.^40,41^ For specific cognitive domains, at risk HDL-C and TC were both associated with worse episodic memory only, which suggests that dysregulation of lipid metabolism might be associated with certain cognitive functions. However, we were not able to identify specific neuropsychiatric or neuropathological mechanisms in this study. We did not observe an association between TC and cognitive status in either ancestry sample, which differs from some existing findings. For example, a prior meta-analysis reported adults with high TC had 2.14 times higher risk of developing dementia compared to those with normal TC.^3^ The difference may be attributed to variations across samples, in that our included study sample captured a lower proportion of abnormal TC participants (**Supplementary Table 1**).

It is worth noticing that in our study of older adults, we only observed significant associations between HDL-C and CIND, but not with dementia. One possible explanation on that is that in our sensitivity analyses, we adjusted for potential mediators, which could be part of the causal chain between the cholesterol level and cognitive impairment. Adjusting for potential mediators can bias those associations.^42^ The attenuated association between HDL-C and CIND after additional adjustments for health conditions also supports this assumption (**Table 6**).

By contrast, the causal inference was only observed for at risk (low) HDL-C levels with dementia. No causal relationships were observed between HDL-C levels and CIND or any cognitive domain scores, including episodic memory. It is possible that PGS for cholesterol capture an accumulation of information across the life course compared to the cross-sectional HDL-C measurements. Given the strong association between HDL PGS and HDL-C levels, a lower HDL PGS could increase the risk of dementia in later life by decreasing HDL-C levels prior to clinical dementia.

To assess the validity of our Mendelian randomization models, several tests were performed to evaluate the assumptions of the testing framework. The relevance assumption is met by providing evidence of both cholesterol PGSs as valid instruments using improvement χ^2^ tests. A limitation of our analysis is the potential violation of the independence and exclusion restriction assumptions. Even though we tested for potential violations of pleiotropy, implemented negative controls, and as a sensitivity analysis used the SNP rs3764261 as a valid instrumental variable, our study cannot rule out the presence of pleiotropy. As such, the inferred causality between HDL-C and cognitive status should be interpreted with caution.

No association was found between cholesterol levels and cognitive status in the African ancestry sample. African ancestry participants are traditionally underrepresented in genetic research. Because the method for computing the cholesterol PGSs depended on summary statistics from GWASs focused exclusively on participants of the European ancestry, results may have limited generalizability to other ancestral groups.^43^ Furthermore, we had a relative small sample of the African ancestry (n_African_ = 2,101 versus n_European_ = 8,781). A similar study should be replicated in an African ancestry with a larger sample size and PGSs based on GWASs among the African ancestry specifically. Previous studies have found that African American men and women have better lipid profiles than their White counterparts, including higher levels of HDL-C, lower levels of LDL-C, TC, and triglycerides (TG).^44,45^ Despite this beneficial lipid profile, African Americans, as a collective group, are at greater risk for chronic conditions that are physiologically related to an unfavourable lipid imbalance, such as: coronary heart disease,^46^ stroke,^47^, and dementia.^3^ In our sample, participants of African ancestry have comparable lipid profiles to those with European ancestry. However, our results indicate that an adverse lipid profile is not predictive of dementia nor CIND among African ancestry participants. Therefore, future research should focus on understanding and identifying clinical biomarkers of cognitive decline for participants with non-European ancestry.

There are several strengths and limitations in our current study. First, studies examining blood cholesterol levels and cognitive function in the context of genetics are limited, and thus our findings contribute to an important, yet sparse, literature. Second, Mendelian randomization has a powerful control for confounding and reverse causation, which often impede or mislead epidemiological studies of causation.^48^ We used summary scores (PGSs) of cumulative variation across multiple genetic loci to capture a larger fraction of variability of blood cholesterol, and thus increased the power of testing. Our findings should be considered in light of potential selection bias. Individuals with dyslipidemia may die prematurely from dyslipidemia-related diseases, such as cardiovascular diseases, before developing dementia. Thus, our sample may have been biased toward healthier individuals – those that survived – which may not be representative in the general older population. In such a case, our estimates are conservative and represent an underestimation of the true causal odds ratio. Furthermore, although the dementia and CIND cut points have been clinically validated with an estimated sensitivity of 78%,^20^ the classification of cognitive status is not as well-defined as other more explicitly defined variables. However, our analysis with multiple domains of cognition suggests that our main finding of a potential causal association between HDL-C and dementia among participants of European ancestry may be robust. We argue that any potential misclassification of dementia by the Langa-Weir cognitive status’ algorithm is non-differential by HDL-C status. Therefore, our estimates are conservative and biased toward the null. Finally, we used concurrent assessments of cholesterol and cognition in our current study, but measurements of early-life cholesterol level were not available. Results of causal tests could be strengthened by further studies with time-specific variables. Similar studies should be replicated with larger sample sizes and in multiple ancestries to promote generalizability.

Dementia is a major public health concern worldwide. Blood cholesterol levels, unlike genetic factors, can be modified by changing diets and health behaviors. Findings in our study underscore the protective effects of increased blood HDL-C and its role in maintaining cognition during aging. Thus, therapeutic strategies aimed at controlling cholesterol levels could be a converging target to mitigate cognitive deficits.

## Data Availability

The Health and Retirement Data are publicly available through the HRS website (https://hrs.isr.umich.edu/about). Raw genetic data on unrelated individuals, both genotyped and imputed to the 1000 Genomes Project, is available from the National Center for Biotechnology Information database of genotypes and phenotypes (dbGaP Study Accession: phs000428.v2.p2).

https://www.ncbi.nlm.nih.gov/gap/

## Acronyms

BMI: body mass index
CI: confidence intervals
CIND: cognitive impairment-non dementia
GWAS: genome-wide association study
HDL-C: high-density lipoprotein cholesterol
HRS: Health and Retirement Study
OR: odds ratios
PC: principal component
PGS: polygenic score
SNP: single nucleotide polymorphisms
TC: total cholesterol

## Declaration of competing financial interests

The authors declare they have no actual or potential competing financial interests.

## Funding and acknowledgements

All authors were supported by grants from the National Institute on Aging (R01 AG055406 and R01 AG067592). Dr. Bakulski was supported by grants from the National Institute for Environmental Health Sciences and the National Institute for Minority Health and Health Disparities (R01 ES025531; R01 ES025574; and R01 MD013299). Dr. Ware was supported by a grant from the National Institute on Aging (R01 AG055654). This work was supported by the Michigan Center for the Demography of Aging (P30 AG012846) and the Michigan Alzheimer’s Disease Center (P30 AG053760). The funders did not have any role in the design, analysis, interpretation, or writing of the manuscript.

## Notes

### Competing Interest Statement

The authors have declared no competing interest.

### Author Declarations

e Health and Retirement Study is sponsored by the National Institute on Aging (NIA U01AG009740) and is conducted by the University of Michigan, where written informed consent was approved by the Institutional Review Board. This analysis was exempt and not regulated as determined by the Institutional Review Board at the University of Michigan (HUM00128220).

## References cited

1. Dementia. Accessed September 27, 2019. https://www.who.int/news-room/fact-sheets/detail/dementia

2. Dementia treatment cost forecast globally 2018-2030. Statista. Accessed July 20, 2020. https://www.statista.com/statistics/471323/global-dementia-economic-impact-forecast/

3. Anstey KJ, Ashby-Mitchell K, Peters R. Updating the Evidence on the Association between Serum Cholesterol and Risk of Late-Life Dementia: Review and Meta-Analysis. J Alzheimers Dis. 2017;56(1):215–228. doi:10.3233/JAD-160826

4. Komulainen P, Lakka TA, Kivipelto M, et al. Metabolic syndrome and cognitive function: a population-based follow-up study in elderly women. Dement Geriatr Cogn Disord. 2007;23(1):29–34. doi:10.1159/000096636

5. Yaffe K, Bahorik AL, Hoang TD, et al. Cardiovascular Risk Factors and Accelerated Cognitive Decline in Midlife: the CARDIA Study. Neurology. Published online July 9, 2020. doi:10.1212/WNL.0000000000010078

6. Koch M, Jensen MK. HDL-cholesterol and apolipoproteins in relation to dementia. Curr Opin Lipidol. 2016;27(1):76–87. doi:10.1097/MOL.0000000000000257

7. M K, El H, Mp L, et al. Apolipoprotein E epsilon4 allele, elevated midlife total cholesterol level, and high midlife systolic blood pressure are independent risk factors for late-life Alzheimer disease. Ann Intern Med. 2002;137(3):149–155. doi:10.7326/0003-4819-137-3-200208060-00006

8. Kathiresan S, Manning AK, Demissie S, et al. Agenome-wide association study for blood lipid phenotypes in the Framingham Heart Study. BMC Med Genet. 2007;8 Suppl 1:S17. doi:10.1186/1471-2350-8-S1-S17

9. Willer CJ, Schmidt EM, Sengupta S, et al. Discovery and refinement of loci associated with lipid levels. Nature Genetics. 2013;45(11):1274–1283. doi:10.1038/ng.2797

10. HRS Data Book | Health and Retirement Study. Accessed December 11, 2019. https://hrs.isr.umich.edu/about/data-book

11. Bullain SS, Corrada MM. Dementia in the oldest old. Continuum (Minneap Minn). 2013;19(2 Dementia):457–469. doi:10.1212/01.CON.0000429172.27815.3f

12. Eileen M. Crimmins, Jessica D Faul, Jung Ki Kim, et al. Documentation of Biomarkers in the 2006 and 2008 Health and Retirement Study. Institute for Social Research, University of Michigan; 2013.

13. Eileen M. Crimmins, Jessica D Faul, Jung Ki Kim, David R Weir. Documentation of Biomarkers in the 2010 and 2012 Health and Retirement Study. Survey Research Center, University of Michigan; 2015.

14. Crimmins E, Kim JK, McCreath H, Faul J, Weir D, Seeman T. Validation of blood-based assays using dried blood spots for use in large population studies. Biodemography Soc Biol. 2014;60(1):38–48. doi:10.1080/19485565.2014.901885

15. Sensitive Health Data | Health and Retirement Study. Accessed December 13, 2019. http://hrsonline.isr.umich.edu/index.php?p=healthdat&_ga=2.253318798.625031607.1576100757-1101446680.1569101058

16. Data Product Detail | Health and Retirement Study. Accessed December 13, 2019. http://hrsonline.isr.umich.edu/index.php?p=shoavail&iyear=XV

17. Grundy SM, Stone NJ, Bailey AL, et al. 2018 AHA/ACC/AACVPR/AAPA/ABC/ACPM/ADA/AGS/APhA/ASPC/NLA/PCNA Guideline on the Management of Blood Cholesterol: A Report of the American College of Cardiology/American Heart Association Task Force on Clinical Practice Guidelines. Circulation. 2019;139(25). doi:10.1161/CIR.0000000000000625

18. Millán J, Pintó X, Muñoz A, et al. Lipoprotein ratios: Physiological significance and clinical usefulness in cardiovascular prevention. Vasc Health Risk Manag. 2009;5:757–765.

19. Ofstedal MB, Fisher G. Documentation of Cognitive Functioning Measures in the Health and Retirement Study. Institute for Social Research, University of Michigan; 2005. doi:10.7826/ISR-UM.06.585031.001.05.0010.2005

20. Crimmins EM, Kim JK, Langa KM, Weir DR. Assessment of Cognition Using Surveys and Neuropsychological Assessment: The Health and Retirement Study and the Aging, Demographics, and Memory Study. J Gerontol B Psychol Sci Soc Sci. 2011;66B(Suppl 1):i162–i171. doi:10.1093/geronb/gbr048

21. Leritz EC, McGlinchey RE, Salat DH, Milberg WP. Elevated Levels of Serum Cholesterol are Associated with Better Performance on Tasks of Episodic Memory. Metab Brain Dis. 2016;31(2):465–473. doi:10.1007/s11011-016-9797-y

22. McArdle JJ, Fisher GG, Kadlec KM. Latent variable analyses of age trends of cognition in the Health and Retirement Study, 1992-2004. Psychology and Aging. 2007;22(3):525–545. doi:10.1037/0882-7974.22.3.525

23. Mary Beth Ofstedal, Gwenith G Fisher, A. Regula Herzog. Documentation of Cognitive Functioning Measures in the Health and Retirement Study. Institute for Social Research, University of Michigan; 2005. http://hrsonline.isr.umich.edu/sitedocs/userg/dr-006.pdf

24. Fisher G, Hassan H, Faul J, Rogers W, Weir D. Health and Retirement Study Imputation of Cognitive Functioning Measures: 1992-2014.

25. Monica 1776 Main Street Santa, California 90401-3208. RAND HRS Longitudinal File 2016 (V1). Accessed October 22, 2019. https://www.rand.org/well-being/social-and-behavioral-policy/centers/aging/dataprod/hrs-data.html

26. David R. W. Quality Control Report for Genotypic Data. Published online September 4, 2013. http://hrsonline.isr.umich.edu/sitedocs/genetics/HRS2_qc_report_SEPT2013.pdf?_ga=2.160262008.498457094.1571249480-1101446680.1569101058

27. Ware EB, Schmitz LL, Gard A, Faul J. HRS Polygenic Scores 2006-2012 Genetic Data - Release 3 | Health and Retirement Study. Accessed October 18, 2019. https://hrs.isr.umich.edu/news/hrs-polygenic-scores-2006-2012-genetic-data-release-3

28. Davies G, Armstrong N, Bis JC, et al. Genetic contributions to variation in general cognitive function: a meta-analysis of genome-wide association studies in the CHARGE consortium (N=53949). Mol Psychiatry. 2015;20(2):183–192. doi:10.1038/mp.2014.188

29. Imputation Report - 1000 Genomes Project reference panel. Published online September 27, 2013.

30. Altman DG, Bland JM. Interaction revisited: the difference between two estimates. BMJ. 2003;326(7382):219. doi:10.1136/bmj.326.7382.219

31. Palmer TM, Sterne JAC, Harbord RM, et al. Instrumental variable estimation of causal risk ratios and causal odds ratios in Mendelian randomization analyses. Am J Epidemiol. 2011;173(12):1392–1403. doi:10.1093/aje/kwr026

32. Hernán MA, Robins JM. Instruments for causal inference: an epidemiologist’s dream?Epidemiology. 2006;17(4):360–372. doi:10.1097/01.ede.0000222409.00878.37

33. Davies NM, Holmes MV, Davey Smith G. Reading Mendelian randomisation studies: a guide, glossary, and checklist for clinicians. BMJ. 2018;362:k601. doi:10.1136/bmj.k601

34. Schierer A, Been LF, Ralhan S, Wander GS, Aston CE, Sanghera DK. Genetic Variation in Cholesterol Ester Transfer Protein (CETP), Serum CETP Activity, and Coronary Artery Disease (CAD) Risk in Asian Indian Diabetic Cohort. Pharmacogenet Genomics. 2012;22(2):95–104. doi:10.1097/FPC.0b013e32834dc9ef

35. R: The R Project for Statistical Computing. Accessed October 18, 2019. https://www.r-project.org/

36. Sparks DL, Kryscio RJ, Connor DJ, et al. Cholesterol and Cognitive Performance in Normal Controls and the Influence of Elective Statin Use after Conversion to Mild Cognitive Impairment: Results in a Clinical Trial Cohort. NDD. 2010;7(1-3):183–186. doi:10.1159/000295660

37. Landi F, Russo A, Cesari M, Pahor M, Bernabei R, Onder G. HDL-cholesterol and physical performance: results from the ageing and longevity study in the sirente geographic area (ilSIRENTE Study). Age Ageing. 2007;36(5):514–520. doi:10.1093/ageing/afm105

38. Holtzman DM. Role of apoe/Abeta interactions in the pathogenesis of Alzheimer’s disease and cerebral amyloid angiopathy. J Mol Neurosci. 2001;17(2):147–155. doi:10.1385/JMN:17:2:147

39. Button EB, Robert J, Caffrey TM, Fan J, Zhao W, Wellington CL. HDL from an Alzheimer’s disease perspective. Current Opinion in Lipidology. 2019;30(3):224–234. doi:10.1097/MOL.0000000000000604

40. Song F, Poljak A, Crawford J, et al. Plasma apolipoprotein levels are associated with cognitive status and decline in a community cohort of older individuals. PLoS ONE. 2012;7(6):e34078. doi:10.1371/journal.pone.0034078

41. Singh-Manoux A, Gimeno D, Kivimaki M, Brunner E, Marmot MG. Low HDL cholesterol is a risk factor for deficit and decline in memory in midlife: the Whitehall II study. Arterioscler Thromb Vasc Biol. 2008;28(8):1556–1562. doi:10.1161/ATVBAHA.108.163998

42. Corraini P, Olsen M, Pedersen L, Dekkers OM, Vandenbroucke JP. Effect modification, interaction and mediation: an overview of theoretical insights for clinical investigators. Clin Epidemiol. 2017;9:331–338. doi:10.2147/CLEP.S129728

43. Martin AR, Gignoux CR, Walters RK, et al. Human Demographic History Impacts Genetic Risk Prediction across Diverse Populations. Am J Hum Genet. 2017;100(4):635–649. doi:10.1016/j.ajhg.2017.03.004

44. Cook CB, Erdman DM, Ryan GJ, et al. The pattern of dyslipidemia among urban African-Americans with type 2 diabetes. Diabetes Care. 2000;23(3):319. doi:10.2337/diacare.23.3.319

45. Cowie CC, Howard BV, Harris MI. Serum lipoproteins in African Americans and whites with non-insulin-dependent diabetes in the US population. Circulation. 1994;90(3):1185–1193. doi:10.1161/01.cir.90.3.1185

46. Leigh JA, Alvarez M, Rodriguez CJ. Ethnic Minorities and Coronary Heart Disease: an Update and Future Directions. Curr Atheroscler Rep. 2016;18(2):9. doi:10.1007/s11883-016-0559-4

47. Kleindorfer Dawn. Sociodemographic Groups at Risk: Race/Ethnicity. Stroke. 2009;40(3_suppl_1):S75–S78. doi:10.1161/STROKEAHA.108.534909

48. Olena Y. Mendelian Ramdomization. Accessed October 19, 2019. https://www.ncbi.nlm.nih.gov/projects/gap/cgi-bin/study.cgi?study_id=phs000428.v2.p2

