## Supplementary Tables and Figures for "Mendelian randomization of dyslipidemia on cognitive impairment among older Americans"

| **Supplementary Figure 1.** Sample selection steps in the Health and Retirement Study sample | |
| --- | --- |
|  | 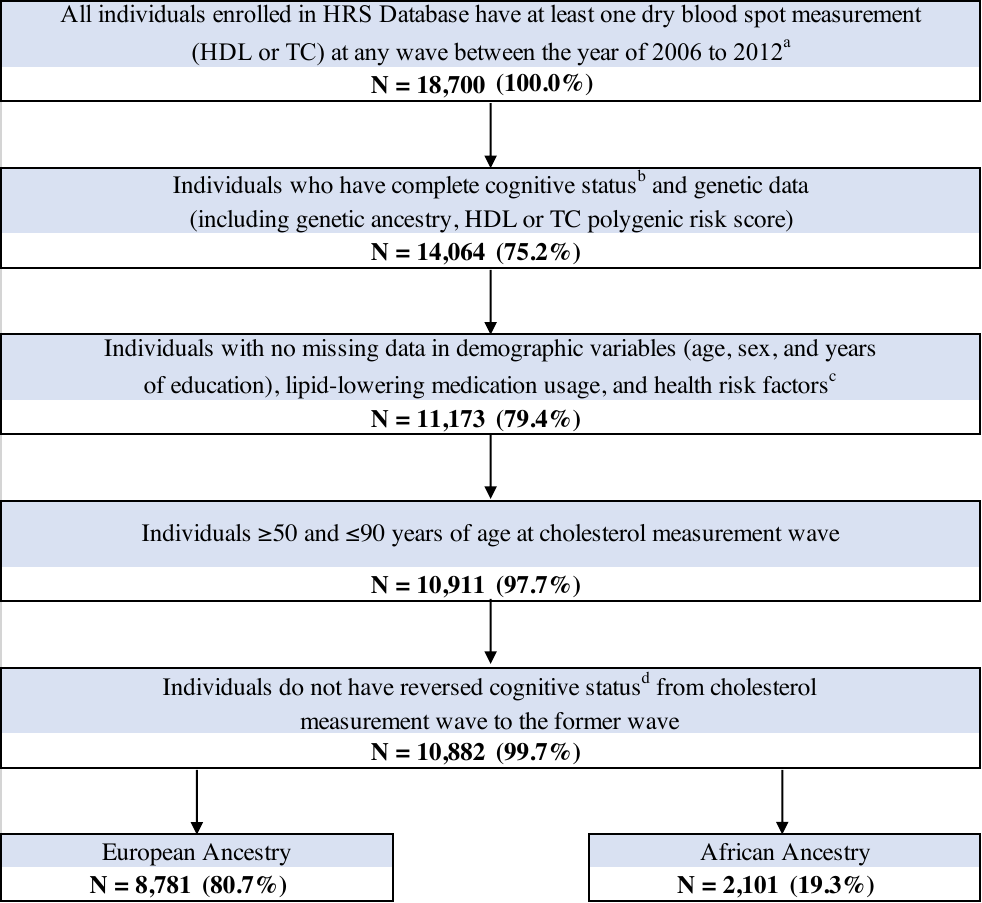 |
| *Abbreviations: HDL, High Density Lipoprotein Cholesterol; TC, Total Cholesterol.* | |
| **Notes:** [a] Samples were identified from the Health and Retirement Study database, which is a national longitudinal panel study of individuals aged over 50 in the United States. Eligible samples should be in at least one wave (e.g. 2006, 2008, 2010, or 2012). [b] Cognitive status was categorized into three levels as normal, cognitive impairment non-dementia, and dementia based on results of a series of cognitive tests. The cut-point for categorization was established by Crimmins et al. [c] Health risk factors including stroke status, body mass index, history of hypertension, diabetes, smoking status, and drink status. [d] Reversed cognition status refers to people classified as normal cognition in current wave (e.g. 2010) but dementia in the former wave (e.g. 2008). | |

| **Supplementary Table 1.** Univariate characteristics of study participants in the Health and Retirement Study sample: included (n = 10,882) vs. excluded sample (n = 7818)^a^ | | | | |
| --- | --- | --- | --- | --- |
|  | **Overall** | **Included** | **Excluded** | **P-value^b^** |
|  | **n = 18,700** | **n = 10,882** | **n = 7818** |  |
| **Categorical Variables [Count (Frequency)]** |  |  |  |  |
| HDL clinical level^c^ |  |  |  | 0.52 |
| Normal | 11583 (68.1%) | 6711 (68.3%) | 4872 (67.8%) |  |
| At risk (low) | 5421 (31.9%) | 3112 (31.7%) | 2309 (32.2%) |  |
| TC clinical level^c^ |  |  |  | <0.001* |
| Normal | 15502 (83.0%) | 9170 (84.4%) | 6332 (81.2%) |  |
| At risk (high) | 3168 (17.0%) | 1700 (15.6%) | 1468 (18.8%) |  |
| Cognitive status at cholesterol measurement wave |  |  |  | <0.001* |
| Normal | 14922 (79.8%) | 8913 (81.9%) | 6009 (76.9%) |  |
| Cognitive Impairment-Non Dementia | 3076 (16.5%) | 1624 (14.9%) | 1452 (18.6%) |  |
| Dementia | 699 (3.74%) | 345 (3.17%) | 354 (4.53%) |  |
| Sex (Female) | 10824 (57.9%) | 6423 (59.0%) | 4401 (56.3%) | <0.001* |
| Gene ancestry (European) | 11314 (80.4%) | 8781 (80.7%) | 2533 (79.6%) | 0.18 |
| Lipid-lowering medication (Yes) | 7401 (49.7%) | 5430 (49.9%) | 1971 (49.2%) | 0.49 |
| Stroke history (Yes) | 1178 (6.30%) | 824 (7.57%) | 354 (4.53%) | <0.001* |
| Hypertension history (Yes) | 10327 (55.3%) | 7150 (65.7%) | 3177 (40.8%) | <0.001* |
| Diabetes history (Yes) | 3875 (20.7%) | 2494 (22.9%) | 1381 (17.7%) | <0.001* |
| Smoking status |  |  |  | <0.001* |
| Never | 7962 (42.8%) | 4554 (41.8%) | 3408 (44.2%) |  |
| Former | 7649 (41.2%) | 4793 (44.0%) | 2856 (37.1%) |  |
| Current | 2976 (16.0%) | 1535 (14.1%) | 1441 (18.7%) |  |
| Drink status (Ever drinker) | 10299 (55.1%) | 5922 (54.4%) | 4377 (56.0%) | 0.03* |
| Proxy status (Self-respondent) | 18697 (100%) | 10882 (100%) | 7815 (100%) | . |
| *APOE-ε4* allele carrier (Yes) | 3899 (27.0%) | 2605 (27.8%) | 1294 (25.5%) | 0.003* |
| Cholesterol measure wave |  |  |  | <0.001* |
| Wave 2006 | 6199 (33.1%) | 3893 (35.8%) | 2306 (29.5%) |  |
| Wave 2008 | 6030 (32.2%) | 3922 (36.0%) | 2108 (27.0%) |  |
| Wave 2010 | 3328 (17.8%) | 1582 (14.5%) | 1746 (22.3%) |  |
| Wave 2012 | 3143 (16.8%) | 1485 (13.6%) | 1658 (21.2%) |  |
| **Continuous Variables [Mean (SD)]** |  |  |  |  |
| Age at cholesterol measurement wave (yrs) | 65.8 (10.8) | 67.6 (10.1) | 63.4 (11.3) | <0.001* |
| Years of education | 12.6 (3.20) | 13.0 (2.63) | 12.0 (3.77) | <0.001* |
| Body Mass Index (kg/m^2^) | 28.6 (6.12) | 28.9 (6.16) | 28.3 (6.04) | <0.001* |
| Immediate word recall | 5.43 (1.60) | 5.50 (1.58) | 5.33 (1.63) | <0.001* |
| Delayed word recall | 4.31 (1.93) | 4.37 (1.92) | 4.23 (1.94) | <0.001* |
| Serial 7 subtraction | 3.44 (1.70) | 3.57 (1.64) | 3.27 (1.75) | <0.001* |
| Backward count from 20 | 1.87 (0.48) | 1.89 (0.45) | 1.85 (0.52) | <0.001* |
| Vocabulary sum score | 5.43 (2.07) | 5.57 (2.04) | 5.24 (2.10) | <0.001* |
| Total cognition | 21.6 (4.99) | 22.1 (4.82) | 20.8 (5.16) | <0.001* |
| *Abbreviations: APOE, Apolipoprotein E; HDL, High Density Lipoprotein Cholesterol; SD: Standard Deviation; TC, Total Cholesterol.* | | | | |
| **Notes:** [a] All variables were measured at the first instance of biomarker collection for a participant from Health and Retirement Study waves 2006-2012. All the statistics including count, frequency, mean, SD, and *P* value were calculated based on non-missing data for each variable. [b] The overall *P* value was calculated from chi-square test or analysis of variance for categorical or continuous variables as appropriate, interpreted as differences between groups. * indicates a significance at a P value of 0.05. [c] At risk low HDL: <40 mg/dL for male and <50 mg/dL for female; At risk high TC: ≥240 mg/dL. | | | | |

| **Supplementary Table 2.** Associations between polygenic risk score for cholesterol and cognitive status, in the Health and Retirement Study, European ancestry sample (n = 8781)^a^ | | | | | | | | |
| --- | --- | --- | --- | --- | --- | --- | --- | --- |
|  |  | **High Density Lipoprotein Cholesterol (HDL)** | | |  | **Total Cholesterol (TC)** | | |
|  |  | **N** | **β coefficient (95% CI)** | |  | **N** | **β coefficient (95% CI)** | |
| **Total effect of cholesterol PGS** |  |  |  |  |  |  |  |  |
| **Total cognition score (0-35)** | Crude | 5930 | 0.100 | (-0.02, 0.22) |  | 5924 | 0.019 | (-0.10, 0.13) |
|  | Adjusted^b^ | 5930 | 0.072 | (-0.03, 0.17) |  | 5924 | 0.036 | (-0.07, 0.15) |
| **Episodic memory** |  |  |  |  |  |  |  |  |
| Immediate word recall | Crude | 8781 | 0.027 | (-0.01, 0.06) |  | 8775 | 0.001 | (-0.03, 0.03) |
|  | Adjusted | 8781 | 0.025 | (-0.00, 0.05) |  | 8775 | 0.009 | (-0.02, 0.04) |
| Delayed word recall | Crude | 8781 | 0.028 | (-0.01, 0.07) |  | 8775 | -0.010 | (-0.05, 0.03) |
|  | Adjusted | 8781 | 0.032 | (-0.00, 0.07) |  | 8775 | 0.009 | (-0.03, 0.05) |
| **Mental status** |  |  |  |  |  |  |  |  |
| Serial 7 subtraction | Crude | 8781 | 0.012 | (-0.02, 0.04) |  | 8775 | 0.010 | (-0.02, 0.04) |
|  | Adjusted | 8781 | -0.005 | (-0.03, 0.02) |  | 8775 | 0.007 | (-0.02, 0.04) |
| Backward count from 20 | Crude | 8781 | 0.001 | (-0.01, 0.01) |  | 8775 | 0.003 | (-0.01, 0.01) |
|  | Adjusted | 8781 | 0.001 | (-0.01, 0.01) |  | 8775 | 0.003 | (-0.01, 0.01) |
| **Vocabulary** |  |  |  |  |  |  |  |  |
| Vocabulary summary | Crude | 3172 | 0.053 | (-0.01, 0.12) |  | 3166 | 0.009 | (-0.06, 0.07) |
| score | Adjusted | 3172 | 0.020 | (-0.04, 0.08) |  | 3166 | -0.002 | (-0.07, 0.06) |
| **Direct effect of cholesterol PGS (adjusting for cholesterol clinical level)** | | | | |  |  |  |  |
| **Total cognition score (0-35)** | Crude | 5270 | 0.080 | (-0.04, 0.20) |  | 5930 | 0.025 | (-0.09, 0.14) |
|  | Adjusted | 5270 | 0.073 | (-0.04, 0.18) |  | 5930 | 0.032 | (-0.08, 0.14) |
| **Episodic memory** |  |  |  |  |  |  |  |  |
| Immediate word recall | Crude | 7869 | 0.020 | (-0.01, 0.05) |  | 8781 | 0.004 | (-0.03, 0.04) |
|  | Adjusted | 7869 | 0.025 | (-0.01, 0.06) |  | 8781 | 0.007 | (-0.02, 0.04) |
| Delayed word recall | Crude | 7869 | 0.015 | (-0.03, 0.06) |  | 8781 | -0.005 | (-0.05, 0.03) |
|  | Adjusted | 7869 | 0.027 | (-0.01, 0.06) |  | 8781 | 0.009 | (-0.03, 0.05) |
| **Mental status** |  |  |  |  |  |  |  |  |
| Serial 7 subtraction | Crude | 7869 | 0.007 | (-0.03, 0.04) |  | 8781 | 0.009 | (-0.02, 0.04) |
|  | Adjusted | 7869 | -0.003 | (-0.03, 0.03) |  | 8781 | 0.005 | (-0.03, 0.04) |
| Backward count from 20 | Crude | 7869 | 0.001 | (-0.01, 0.01) |  | 8781 | 0.003 | (-0.01, 0.01) |
|  | Adjusted | 7869 | 0.002 | (-0.01, 0.01) |  | 8781 | 0.003 | (-0.01, 0.01) |
| **Vocabulary** |  |  |  |  |  |  |  |  |
| Vocabulary summary | Crude | 2714 | 0.064 | (-0.01, 0.14) |  | 3172 | 0.010 | (-0.06, 0.07) |
| score | Adjusted | 2714 | 0.035 | (-0.03, 0.10) |  | 3172 | -0.003 | (-0.07, 0.06) |
| *Abbreviations: CI, Confidence Interval; CIND, Cognitive Impairment-Non Dementia; OR, Odds Ratio; PGS, Polygenic Score* | | | | | | | | |
| **Notes:** [a] All variables were measured at the first instance of biomarker collection for a participant from Health and Retirement Study waves 2006-2012. All the values were based on results from linear regression analyses in each sample.  [b] Adjusted for age, sex, years of education, lipid-lowering medication, cholesterol measurement wave, and five ancestry-specific principal components. | | | | | | | | |

| **Supplementary Table 3.** Associations and causal inference between rs3764261, HDL-C level, and cognitive status, Health and Retirement Study, European ancestry sample (n = 8781)^a^ | | | | | | | | |
| --- | --- | --- | --- | --- | --- | --- | --- | --- |
|  | **Overall sample** | |  | **CIND vs. Normal** | |  | **Dementia vs. Normal** | |
|  | **n = 6491** | |  | **n = 6350** | |  | **n = 5704** | |
|  | **OR** | **(95 CI%)** |  | **OR** | **(95 CI%)** |  | **OR** | **(95 CI%)** |
| **Model 1: HDL-C level ~ rs3764261** | | | | | | | | |
| Crude | 0.80 | (0.74, 0.86) | | 0.80 | (0.74, 0.87) |  | 0.79 | (0.73, 0.86) |
| Adjusted^b^ | 0.80 | (0.73, 0.86) | | 0.80 | (0.74, 0.87) |  | 0.79 | (0.72, 0.86) |
| **Improvement ꭓ^2 c^** | 1669.0 | |  | 1643.1 | |  | 1470.5 | |
| **Model 2: Cognitive status ~ rs3764261 (+ HDL-C level)** | | | | | | | | |
| **Total effect of rs3764261** | |  |  |  |  |  |  |  |
| Crude |  |  |  | 1.06 | (0.94, 1.18) |  | 1.01 | (0.79, 1.30) |
| Adjusted^b^ |  |  |  | 1.05 | (0.93, 1.18) |  | 0.99 | (0.76, 1.28) |
| **Direct effect of rs3764261 (adjusting for HDL-C level)** | | | | | |  |  |  |
| Crude |  |  |  | 1.07 | (0.96, 1.20) |  | 1.03 | (0.80, 1.32) |
| Adjusted^b^ |  |  |  | 1.06 | (0.94, 1.19) |  | 0.99 | (0.76, 1.28) |
| **Model 3: Mendelian randomization (Wald-type/ratio): Cognitive status ~ HDL-C level** | | | | | | | | |
| Crude |  |  |  | 0.78 | (0.48, 1.29) |  | 0.94 | (0.32, 2.74) |
| Adjusted^b^ |  |  |  | 0.81 | (0.48, 1.37) |  | 1.04 | (0.35, 3.11) |
| *Abbreviations: CI, Confidence Interval; CIND, Cognitive Impairment-Non Dementia; HDL-C, High Density Lipoprotein Cholesterol; OR: Odds Ratio* | | | | | | | | |
| **Notes:** [a] All variables were measured at the first instance of biomarker collection for a participant from Health and Retirement Study waves 2006-2012. All the values were based on results from multivariable logistic regression analyses or mendelian randomization analysis in each sample, in which "normal cognitive status" and "normal cholesterol level" were used as the reference groups.  [b] Adjusted for age, sex, years of education, lipid-lowering medication, cholesterol measurement wave, and five ancestry-specific principal components. [c] Calculated by 2*(log likelihood of full model - log likelihood of reduced model). Statistics larger than 10 indicates a valid instrument in convention. | | | | | | | | |

| **Supplementary Table 4.** Associations between polygenic risk score for cholesterol (HDL-C and TC) and cognitive status in normal cholesterol level sample(s), in the Health and Retirement Study, European ancestry sample^a^ | | | | | | | | | | | | | | | |
| --- | --- | --- | --- | --- | --- | --- | --- | --- | --- | --- | --- | --- | --- | --- | --- |
|  | **CIND vs. Normal** | | | | | | |  | **Dementia vs. Normal** | | | | | | |
|  | **HDL-C** | | |  | **TC** | | |  | **HDL-C** | | |  | **TC** | | |
|  | **N** | **OR** | **(95 CI%)** |  | **N** | **OR** | **(95 CI%)** |  | **N** | **OR** | **(95 CI%)** |  | **N** | **OR** | **(95 CI%)** |
| **Total effect of cholesterol PGS** |  |  |  |  |  |  |  |  |  |  |  |  |  |  |  |
| Crude | 2715 | 0.98 | (-0.15, 0.11) |  | 3330 | 0.96 | (-0.15, 0.07) |  | 2503 | 0.84 | (-0.45, 0.10) |  | 3038 | 0.90 | (-0.34, 0.13) |
| Adjusted (demographic)^b^ | 2715 | 0.95 | (-0.19, 0.09) |  | 3330 | 0.97 | (-0.15, 0.10) |  | 2503 | 0.87 | (-0.42, 0.14) |  | 3038 | 0.89 | (-0.38, 0.14) |
| Adjusted (health status)^c^ | 2715 | 0.96 | (-0.18, 0.10) |  | 3330 | 0.97 | (-0.16, 0.10) |  | 2503 | 0.86 | (-0.45, 0.13) |  | 3038 | 0.91 | (-0.37, 0.17) |
| *Abbreviations: APOE, Apolipoprotein E; CI, Confidence Interval; CIND, Cognitive Impairment-Non Dementia; HDL, High Density Lipoprotein Cholesterol; OR, Odds Ratio; PGS, Polygenic Score; TC, Total Cholesterol* | | | | | | | | | | | | | | | |
| **Notes:** [a] All variables were measured at the first instance of biomarker collection for a participant from Health and Retirement Study waves 2006-2012. All the values were based on results from multivariable logistic regression analyses in each sample, in which "normal cognitive status" and "normal cholesterol level" were used as the reference groups.  [b] Adjusted for age, sex, years of education, lipid-lowering medication, cholesterol measurement wave, and five ancestry-specific principal components. [c] Adjusted for ever drink alcohol, history of stroke, hypertension, diabetes, and BMI in addition to variables in [b]. | | | | | | | | | | | | | | | |

| **Supplementary Table 5.** Associations between TC/HDL cholesterol ratio (log-transformed) and cognitive status in the Health and Retirement Study, European ancestry sample (n = 8781)^a^ | | | | | | | |
| --- | --- | --- | --- | --- | --- | --- | --- |
|  | **CIND vs. Normal** | | |  | **Dementia vs. Normal** | | |
|  | **N** | **OR** | **(95 CI%)** |  | **N** | **OR** | **(95 CI%)** |
| Crude | 7701 | 1.15 | (0.91, 1.47) |  | 6920 | 1.24 | (0.71, 2.14) |
| Adjusted (demographic)^b^ | 7701 | 1.17 | (0.90, 1.53) |  | 6920 | 0.97 | (0.53, 1.77) |
| Adjusted (health status)^c^ | 7701 | 1.11 | (0.84, 1.46) |  | 6920 | 1.00 | (0.53, 1.87) |
| *Abbreviations: APOE, Apolipoprotein E; BMI, Body Mass Index; CI, Confidence Interval; CIND, Cognitive Impairment-Non Dementia; HDL, High Density Lipoprotein Cholesterol; OR, Odds Ratio; TC, Total Cholesterol* | | | | | | | |
| **Notes:** [a] All variables were measured at the first instance of biomarker collection for a participant from Health and Retirement Study waves 2006-2012. All the values were based on results from multivariable logistic regression analyses in each sample, in which "normal cognitive status" was used as the reference group.  [b] Adjusted for age, sex, years of education, lipid-lowering medication, cholesterol measurement wave, and five ancestry-specific principal components. [c] Adjusted for ever drink alcohol, history of stroke, hypertension, diabetes, and BMI in addition to variables in [b]. | | | | | | | |

| **Supplementary Figure 2.** Non-linear associations between continuous cholesterol and cognitive status, in the Health and Retirement Study, European ancestry sample (n = 8781)^a^ | | |
| --- | --- | --- |
| **HDL** | 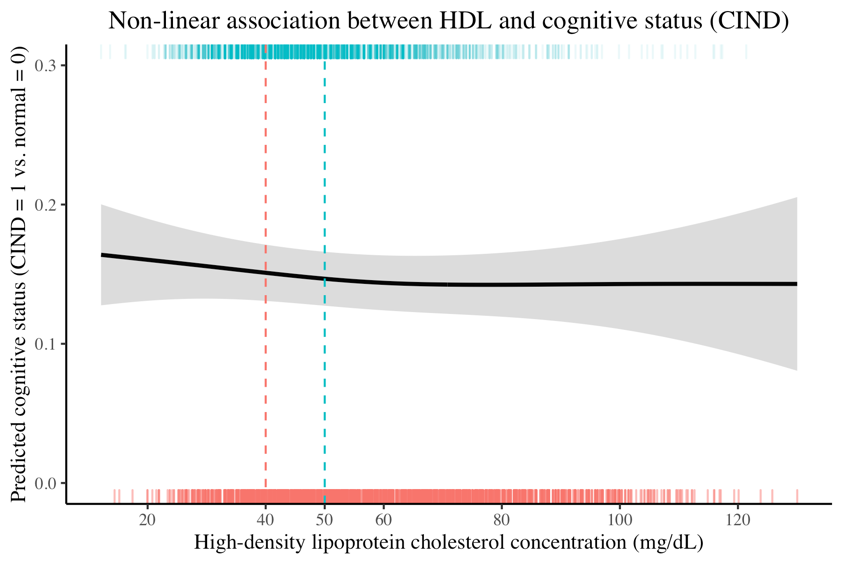 | 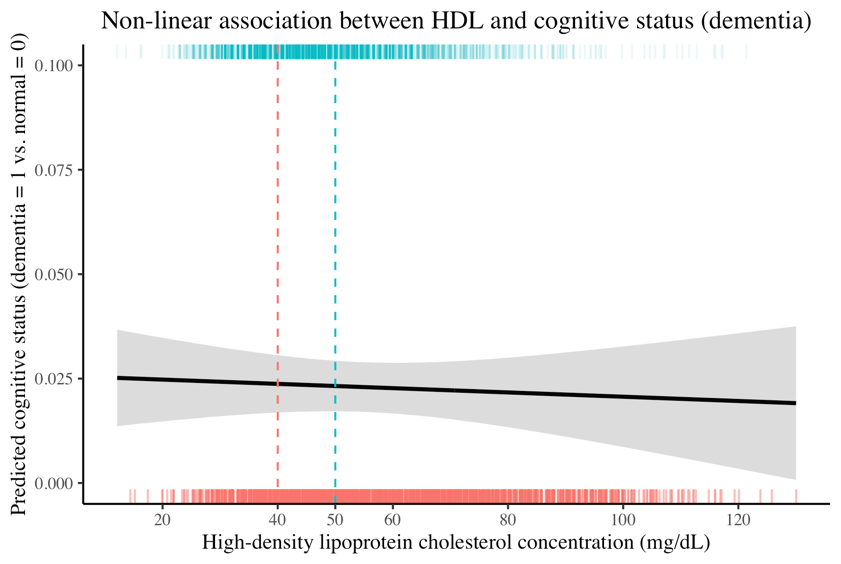 |
| **TC** | 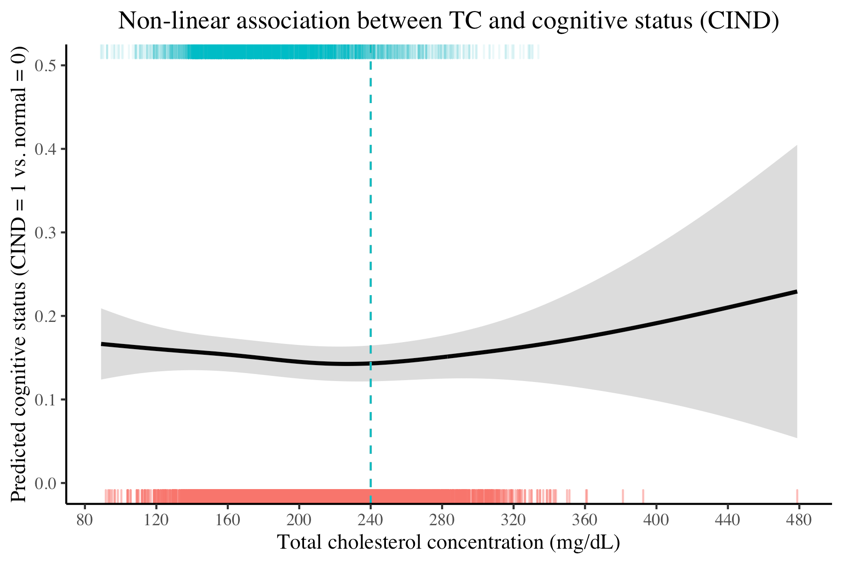 | 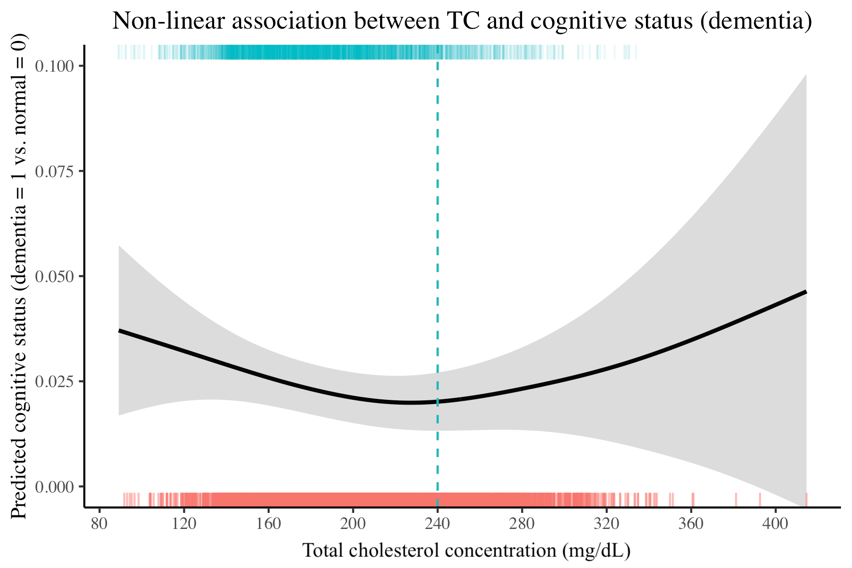 |
| *Abbreviations: CI, Confidence Interval; CIND, Cognitive Impairment-Non Dementia; HDL, High Density Lipoprotein Cholesterol; TC, Total Cholesterol* | | |
| **Notes:** [a] All variables were measured at the first instance of biomarker collection for a participant from Health and Retirement Study waves 2006-2012. All the values were based on results from multivariable logistic regression analyses in each sample, in which "normal cognitive status" was used as the reference group. In the regression model, the continuous cholesterol variable was treated as a spline term; x-axis represents the concentration of cholesterol (mg/dL) (red: female, green: male) and y-axis represents the predicted cognitive status; shaded areas represents confidence intervals for spline terms; dotted lines refer to clinical cutoffs for at risk/normal. Models were adjusted for age, sex, years of education, lipid-lowering medication, cholesterol measurement wave, and five ancestry-specific principal components. | | |

| **Supplementary Table 6a.** Associations between polygenic risk score for cholesterol (HDL-C and TC) and blood cholesterol levels, in the Health and Retirement Study, African ancestry sample (n = 2101)^a^ | | | | | | | | | | | |
| --- | --- | --- | --- | --- | --- | --- | --- | --- | --- | --- | --- |
|  | **CIND & Normal** | | |  | **Dementia & Normal** | | |  | **Overall sample** | | |
|  | **N** | **OR** | **(95 CI%)** |  | **N** | **OR** | **(95 CI%)** |  | **N** | **OR** | **(95 CI%)** |
| **High Density Lipoprotein Cholesterol (HDL-C)** | | | |  |  |  |  |  |  |  |  |
| Crude | 1818 | 0.91 | (0.82, 1.00) |  | 1429 | 0.87 | (0.77, 0.97) |  | 1956 | 0.88 | (0.80, 0.97) |
| Adjusted^b^ | 1818 | 0.93 | (0.83, 1.03) |  | 1429 | 0.87 | (0.77, 0.98) |  | 1956 | 0.89 | (0.81, 0.99) |
| **Improvement ꭓ^2 c^** | 2.08 | | |  | 5.40 | | |  | 4.96 | | |
| **Total Cholesterol (TC)** |  |  |  |  |  |  |  |  |  |  |  |
| Crude | 1935 | 1.14 | (1.01, 1.29) |  | 1532 | 1.17 | (1.02, 1.34) |  | 2095 | 1.16 | (1.03, 1.30) |
| Adjusted | 1935 | 1.18 | (1.04, 1.33) |  | 1532 | 1.22 | (1.06, 1.41) |  | 2095 | 1.20 | (1.07, 1.35) |
| **Improvement ꭓ^2^** | 6.91 | | |  | 7.64 | | |  | 9.09 | | |
| *Abbreviations: CI, Confidence Interval; CIND, Cognitive Impairment-Non Dementia; HDL, High Density Lipoprotein Cholesterol; OR: Odds Ratio; TC, Total Cholesterol* | | | | | | | | | | | |
| **Notes:** [a] All variables were measured at the first instance of biomarker collection for a participant from Health and Retirement Study waves 2006-2012. All the values were based on results from multivariable logistic regression analyses in each sample, in which "normal cholesterol level" was used as the reference group.  [b] Adjusted for age, sex, years of education, lipid-lowering medication, cholesterol measurement wave, and five ancestry-specific principal components. [c] Calculated by 2*(log likelihood of full model - log likelihood of reduced model). Statistics larger than 10 indicates a valid instrument in convention. | | | | | | | | | | | |

| **Supplementary Table 6b.** Associations between polygenic risk for cholesterol (HDL-C and TC), blood cholesterol levels, and cognitive status, in the Health and Retirement Study, African ancestry sample (n = 2101)^a^ | | | | | | | | | |
| --- | --- | --- | --- | --- | --- | --- | --- | --- | --- |
|  | **High Density Lipoprotein Cholesterol (HDL-C)** | | | |  | **Total Cholesterol (TC)** | | | |
|  | **CIND vs. Normal** | | **Dementia vs. Normal** | |  | **CIND vs. Normal** | | **Dementia vs. Normal** | |
|  | **n = 1818** | | **n = 1429** | |  | **n = 1935** | | **n = 1532** | |
|  | **OR** | **(95 CI%)** | **OR** | **(95 CI%)** |  | **OR** | **(95 CI%)** | **OR** | **(95 CI%)** |
| **Model 1: Cognitive Status ~ cholesterol PGS** | | |  |  |  |  |  |  |  |
| **Total effect of cholesterol PGS** | | |  |  |  |  |  |  |  |
| Crude | 0.95 | (0.86, 1.05) | 1.03 | (0.86, 1.23) |  | 1.10 | (1.00, 1.21) | 1.13 | (0.96, 1.34) |
| Adjusted^b^ | 1.00 | (0.90, 1.12) | 1.10 | (0.89, 1.36) |  | 1.09 | (0.98, 1.21) | 1.05 | (0.86, 1.29) |
| **Direct effect of cholesterol PGS (adjusting for cholesterol clinical level)** | | | | | | | |  |  |
| Crude | 0.95 | (0.86, 1.05) | 1.02 | (0.86, 1.22) |  | 1.10 | (1.00, 1.21) | 1.14 | (0.97, 1.35) |
| Adjusted | 1.00 | (0.90, 1.12) | 1.10 | (0.89, 1.36) |  | 1.09 | (0.98, 1.21) | 1.06 | (0.86, 1.30) |
| **Model 2: Cognitive Status ~ cholesterol level** | | |  |  |  |  |  |  |  |
| Crude | 0.98 | (0.79, 1.22) | 0.90 | (0.60, 1.31) |  | 0.89 | (0.68, 1.16) | 0.70 | (0.42, 1.11) |
| Adjusted | 0.99 | (0.78, 1.26) | 0.89 | (0.55, 1.42) |  | 0.99 | (0.74, 1.32) | 0.84 | (0.45, 1.49) |
| *Abbreviations: CI, Confidence Interval; CIND, Cognitive Impairment-Non Dementia; OR, Odds Ratio; PGS, Polygenic Score* | | | | | | | | | |
| **Notes:** [a] All variables were measured at the first instance of biomarker collection for a participant from Health and Retirement Study waves 2006-2012. All the values were based on results from multivariable logistic regression analyses in each sample, in which "normal cognitive status" and "normal cholesterol level" were used as the reference groups.  [b] Adjusted for age, sex, years of education, lipid-lowering medication, cholesterol measurement wave, and five ancestry-specific principal components. | | | | | | | | | |
